# Rule learning in adolescents at clinical high risk for psychosis shows altered patterns of activation

**DOI:** 10.1101/2020.01.20.20018069

**Authors:** Joseph M. Orr, Jesus Lopez, Michael J. Imburgio, Andrea Pelletier-Baldeli, Jessica A. Bernard, Vijay A. Mittal

**Affiliations:** Department of Psychological and Brain Sciences, Texas A&M University, College Station, Texas; Texas A&M Institute for Preclinical Studies, Texas A&M University, College Station, Texas; Department of Psychiatry, UNC-Chapel Hill School of Medicine, Chapel Hill, North Carolina; Department of Psychiatry, Northwestern University, Evanston, Illinois; Medical Social Sciences, Northwestern University, Evanston, Illinois; Institute for Policy Research (IPR), Northwestern University, Evanston, Illinois; Institute for Innovations in Developmental Sciences (DevSci), Northwestern University, Evanston, Illinois

**Keywords:** high risk, psychosis, fMRI, executive function, rule learning, imaging

## Abstract

**Background:** The ability to flexibly apply rules to novel situations is a critical aspect of adaptive human behavior. While executive function deficits are known to appear early in the course of psychosis, it is unclear which specific facets are affected. Identifying whether rule learning is impacted at the early stages of psychosis is necessary for truly understanding the etiology of psychosis and may be critical for designing novel treatments. Therefore, we examined rule learning in healthy adolescents and those meeting criteria for clinical high risk (CHR) for psychosis.

**Methods:** 24 control and 22 CHR adolescents underwent rapid, high-resolution fMRI while performing a paradigm which required them to apply novel or practiced task rules.

**Results:** Previous work has suggested that practiced rules rely on rostrolateral prefrontal cortex (RLPFC) during rule encoding and dorsolateral prefrontal cortex (DLPFC) during task performance, while novel rules show the opposite pattern. We failed to replicate this finding, with greater activity for novel rules during performance. Comparing the HC and CHR group, there were no statistically significant effects, but an effect size analysis found that the CHR group showed less activation during encoding and greater activation during performance. This suggests the CHR group may use less efficient reactive control to retrieve task rules at the time of task performance, rather than proactively during rule encoding.

**Conclusions:** These findings suggest that flexibility may be altered in the clinical high risk state, however, more data is needed to determine whether these deficits predict disease progression.

## Introduction

Unlike most laboratory animals that require extensive training to acquire a task, humans have the ability to rapidly acquire new tasks based on limited instructions. This ability, a critical component of the broader construct of executive function, is critical in dynamic environments where one must adapt practiced knowledge to new instructions (Braver and Barch, 2006; Monsell, 1996; Woolgar et al., 2011). It is generally accepted that these executive function processes are coordinated by the frontal lobes (Duncan, 2010, 1986; 2001; Stuss and Alexander, 2000). Dysfunction of the frontal lobes has been widely described in schizophrenia, from studies of task-related functional activation (MacDonald and Carter, 2003; Minzenberg et al., 2009; Poppe et al., 2016), resting-state functional connectivity (Pettersson-Yeo et al., 2011; Repovs et al., 2011; Rotarska-Jagiela et al., 2010; Zhou et al., 2007), white matter connectivity (Camchong et al., 2009; Oh et al., 2009; van den Heuvel et al., 2010), and structural morphometry (Gur et al., 2000; Sallet et al., 2003). Moreover, it has been suggested that dysfunction of the dorsolateral prefrontal cortex (DLPFC) is related to deficits in the ability to maintain task rules and goals, a core deficit of schizophrenia. Increasing evidence suggests that executive function deficits and prefrontal dysfunctions are present at the prodromal or risk stage of schizophrenia (Allen et al., 2012; Fornito et al., 2013; Fusar-Poli et al., 2011; Harris et al., 2004; Morey et al., 2005; Seidman et al., 2006; Stanfield et al., 2008).

The onset of psychosis is usually preceded by a prodromal phase characterized by functional decline and subtle attenuated symptoms that include positive phenomena and a decline in socio-occupational functioning (Yung and McGorry, 1996). Those at clinical high-risk (CHR; i.e, meeting criteria for a psychosis risk syndrome) are of critical importance as the prodromal period is of interest both as a window for investigating processes involved in disease onset, and also as a potential point of intervention and prevention (Haroun et al., 2006; McGlashan et al., 2007; Thompson et al., 2015). More specifically, recent studies have suggested that adolescents with a prodromal syndrome (i.e., showing moderate attenuated positive symptoms accompanied by a global decline in functioning) (Miller et al., 1999; Woods et al., 2014) are at imminent risk for conversion to a psychotic disorder; although successful early identification and other factors relating to heterogeneous assessment/inclusion criteria have yielded a global decrease in transition rates (Fusar-Poli et al., 2016; Yung et al., 2007), a substantial proportion (anywhere from 10-35%) will convert to a psychotic disorder within a two-year period (Cannon et al., 2016, 2008; Yung et al., 2007).

While cognitive impairments have been well documented in psychosis risk, including in broad domains of executive function (Bora and Murray, 2014; Carrión et al., 2018; Fusar-Poli et al., 2012), these deficits have largely been demonstrated with traditional neuropsychological assessments. This makes it difficult to determine what specific executive function deficits are present in the high-risk period of psychosis. Recently, Guo and colleagues (Guo et al., 2019) examined whether performance on the AX variant of the Continuous Performance Task (AX-CPT)—a task thought to measure context or goal maintenance—predicted progression (i.e., conversion risk) in an at-risk population. They found that baseline performance on the AX-CPT was predictive of clinical status 12 months later. This study confirms earlier proposals of context/goal maintenance as a marker of psychosis risk (Niendam et al., 2014), and is in line with the suggestion that goal maintenance is a core deficit of schizophrenia (Barch and Ceaser, 2012).

While adaptive executive function has been extensively studied in behavioral and neuroimaging studies, these studies have largely relied on highly practiced tasks. Dumontheil and colleagues (2011) demonstrated that new rules are encoded across a broad network of frontal and parietal regions that Duncan (2010) has referred to as the Multiple Demand Network. As tasks become more difficult (but not necessarily more abstract or complex), more rostral regions of the frontal lobes come online (2012). However, several studies on rapid instructed task learning (RITL) by Cole and colleagues (2017, 2016, 2010) have shown that practiced and novel tasks rely on the same regions of the lateral frontal cortex, but the temporal dynamics of these regions varies based on novelty. They demonstrated that practiced task rule encoding relies on the rostrolateral prefrontal cortex (RLPFC) for retrieving task rules from long-term memory, and subsequent rule activation by the dorsolateral prefrontal cortex (DLPFC) for task performance. Novel task preparation showed a reversal of these dynamics, such that the novel rules are encoding in a bottom-up fashion by the DLPFC and become integrated by the RLPFC during task performance. However, there have been no investigations of whether psychosis risk is associated with deficits in learning new rules/tasks In order to better understand which executive function processes are impaired among those at high-risk for psychosis, and map the affected underlying neurobiology, we investigated RITL in CHR adolescents and healthy controls (HC). The paradigm was adapted from the Permuted Rule Operations task of Cole and colleagues (2010), with timing modified slightly for fast multiband fMRI. Participants were extensively trained on 4 combinations of rules about a week before scanning. During scanning, participants saw these same 4 practiced rules, as well 60 novel rule combinations. Given the evidence discussed above that goal maintenance, supported by activity in the DLPFC, is impacted across the psychosis spectrum (MacDonald et al., 2005; Niendam et al., 2014; Poppe et al., 2016), we predicted that CHR participants would show decreased DLPFC activation during novel task rule encoding. Furthermore, in line with the idea that the DLPFC is critical for task rule encoding, we predicted that practiced tasks would be associated with decreased RLPFC activation during encoding and decreased DLPFC activation during performance. Within control participants, we expected to replicate the DLPFC-RLPFC dynamics previously demonstrated by Cole and colleagues (2010).

## Methods

### Participants

Here, we investigated 23 adolescent and young adult CHR participants (mean age= 20.8 + 1.54 years, 7 female), and 25 HC participants (mean age = 21.5 + 1.83 years, 11 female). All participants had previously enrolled in a longitudinal study investigating psychosis risk as part of the Adolescent Development and Preventative Treatment (ADAPT) research program at the University of Colorado Boulder. Participants were recruited for participation in this investigation at the end of their annual study visit, or were directly contacted over the phone. In addition to the current procedures, participants also completed 2 other short paradigms in the same scanning session (Damme et al., 2019; Pelletier-Baldelli et al., 2018). Prior to participating in the imaging study, all participants were consented specifically for the imaging study, and declining to participate did not affect their participation in the ongoing longitudinal study. All procedures were reviewed and approved by the University of Colorado Boulder Institutional Review Board.

Exclusion criteria for both groups included a history of head injury, the presence of a neurological disorder, life-time substance dependence as assessed by the Structured Clinical Interview for Axis-I DSM IV Disorders (First et al., 1995), and the presence of any contraindications for the magnetic resonance imaging environment. In the CHR group, we also excluded individuals with an Axis I psychotic disorder. In the control sample, we excluded individuals with any diagnosis of an Axis I disorder. Further, the presence of a psychotic disorder in first-degree relatives was an additional exclusion criterion for the control group. Due to response box errors (1 CHR participant) and a failure to follow task instructions (1 HC participant), the final sample included 46 participants. See Table 1 for demographics and symptom information.

**Table 1.** Symptoms and demographics

| Table 1. Symptoms and demographics |  |  |  |  |
| --- | --- | --- | --- | --- |
|  | **CHR**<br>*M(SEM)* | **HC**<br>*M(SEM)* | **Statistic** | ***p*** |
| Age | 20.8(1.54) | 21.5(1.83) | $t(44) = 2.23$ | 0.05 |
| Education (years) | 13.25(0.24) | 13.55(0.34) | $t(44) = 0.73$ | 0.47 |
| Parent education (years) | 16.21(0.48) | 15.60(0.62) | $t(44) = 0.78$ | 0.44 |
| Total positive symptoms (SIPS) | 12.23(4.19) | 6.35(6.49) | $t(44) = 3.52$ | 0.001 |
| Gender | CHR (n) | HC (n) | $\chi^2(1) = 0.83$ | 0.36 |
| Male | 13 | 14 |  |  |
| Female | 9 | 11 |  |  |
| Total | 22 | 24 |  |  |
| Abbreviations: CHR, clinical high risk; HC, healthy control; SIPS, Structured Interview for Prodromal Syndromes. |  |  |  |  |

### Symptom Assessment

The Structured Interview for Prodromal Syndromes (SIPS) measures distinct categories of prodromal symptom domains (positive, negative, disorganized, general) and is scored from 0-6 for each symptom. Inclusion in the CHR group was determined by moderate levels of positive symptoms (a SIPS score of 3-5 in one or more of the 5 positive symptom categories), and/or a decline in global functioning in association with the presence of schizotypal personality disorder, and/or a family history of schizophrenia (Miller et al., 1999). All interviewers had inter-rater reliabilities that exceeded Kappa ≥ 80. We confirmed CHR diagnosis for those who participated more than 1 month from entry to larger CHR study protocols. All CHR participants were not taking antipsychotics at the time of participation.

### Permuted Rule Operations (PRO) Paradigm

The Permuted Rule Operations (PRO) Paradigm was adapted from E-Prime code kindly provided by Michael W. Cole and is described in full detail elsewhere (Cole et al., 2010). This paradigm combines 3 types of cues (logic cue, semantic cue, and response cue), each with 4 possibilities to yield 64 possible rule- or task-sets that describe how the participant was to respond to a set of 3 pairs of trial stimuli (See Figure 1). The trial stimuli consisted of concrete nouns and a participant’s task was to indicate whether the stimuli were True or False with respect to the rule. The response cue indicated which button was to be used to indicate True, and the other possible finger (index or middle) of the same hand was to be used to indicate False. The same rule applied to all three trials in a block. For example, if the set of cues was SAME (logic cue), SWEET (semantic cue), and LEFT INDEX (response cue), and the trial stimuli were SEAWEED + TURNIP, GRAPE + APPLE, and FUR + SUGAR, a participant would respond TRUE (both are not sweet), TRUE (both are sweet), FALSE (one is not sweet, the other is sweet). At the beginning of an experimental block the cues were presented one at a time, each for 0.92s (2 TR). After a variable delay between 1.84s and 5.98s (4-6 TR), participants performed 3 trials. On each trial, the two stimuli were presented one at a time, each for 0.92s (2 TR); participants were instructed to respond after the second stimuli was presented. There was a variable inter-trial delay between 1.84s and 5.98s (4-12 TR), and a variable inter-block interval between 11.96s and 16.1s (24-36 TR).

**Figure 1.**
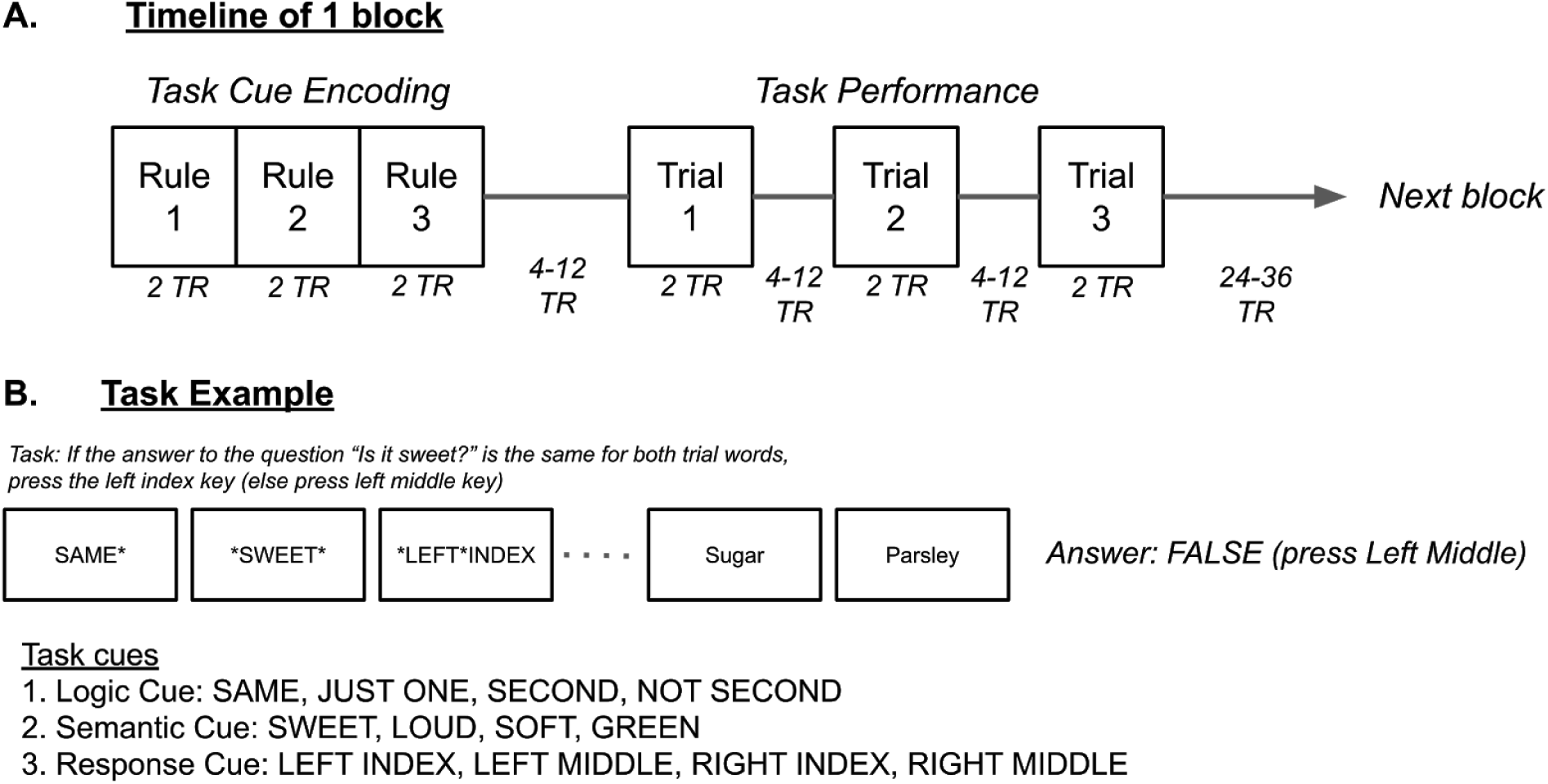
A. Example task block. At the beginning of a block three instruction cues were presented that defined the current task. Each cue was presented one at a time in the following order: logic cue, semantic cue, response cue. There were 4 possible logic cues, 4 possible semantic cues, and 4 possible response cues, yielding 64 possible tasks from all combinations. Of the 64 possible tasks, 4 were practiced before scanning and the remainder were only shown once each in a scanning session. Participants then performed three trials of the task, indicating if the current task rule was true or false. B. An example of one possible task, with the task cues and one trial. Given the cues SAME**SWEET**LEFT*INDEX and the trial stimuli Sugar + Parsley, the answer would be FALSE, as sugar is sweet, but parsley is not.

Of these 64 possible rules, 4 were randomly selected to be practiced during a pre-scan training session which occurred about a week prior to the scan; the practiced rules were counterbalanced across participants. During training, participants received extensive instruction on how to apply the rules, with self-paced examples. Once they understood the instructions, participants completed 12 runs of training, each consisting of 12 blocks of trials with each of the 4 rules being presented 3 times. The first 2 runs were self-paced practice with feedback. After practice, they completed 10 runs without feedback with the same timing as the scanner. They were given time to rest in between blocks. During scanning, participants performed 6 runs consisting of 6 novel and 6 practiced rule blocks. Each novel rule was only presented once in a session, so that not all 64 rules were seen by all participants.

Behavioral data from the training session and the scanning session were analyzed using *jamovi (v. 1*.*0, The jamovi project, 2019)*, a free software package that runs on R. Only correct reaction times were analyzed, and reaction time data and accuracy data were checked for violations of normality with Shapiro-Wilk tests. When normality was violated, non-parametric tests were used.

### Data Sharing

Behavioral data and analysis scripts are available on Open Science Framework (https://osf.io/snuqj/). Raw imaging data are available on OpenNeuro (doi://10.18112/openneuro.ds001371.v1.0.0). Final statistical results are available on BALSA (https://balsa.wustl.edu/Klv19). Additional information about symptoms and demographics are not publicly available, but can be made available by contacting author VAM.

### fMRI Data Acquisition

All functional imaging data was collected using a 3T Siemens Magnetom Tim Trio (software version VB17A; Munich, Germany), using multi-band functional pulse sequences with a 32-channel head coil. Sequences for multi-band functional imaging were acquired from the Center for Magnetic Resonance Research (http://www.cmrr.umn.edu/multiband/index.shtml) and modified as needed for the UCB scanner. Structural images were acquired using a sagittal T1-weighted interleaved sequence (repetition time (TR) = 2400 ms, echo-time (TE) = 2.01 ms, echo spacing = 7.4 ms, flip angle = 8°, field-of-view = 256 mm × 256 mm × 180 mm, voxel resolution = 0.8 mm isotropic). Six runs of multiband EPIs were acquired in the posterior to anterior direction with the following parameters (multiband acceleration factor = 8, bandwidth = 2772 Hz/Px, TR = 460 ms, TE = 29.0, echo-spacing = 0.51 ms, flip-angle = 44°, field-of-view = 248 × 248 × 168 mm, voxel resolution = 3.0 mm isotropic, number of slices = 56, time = 4:00 minutes). We also collected two brief (2 volumes each) scans prior to each of the functional imaging runs, using the same EPI parameters but collected in both the anterior-to-posterior and posterior-to-anterior directions. These scans acquired in order to estimate and correct for distortion (Andersson et al., 2003). The 6 runs of functional data were collected while individuals were performing the PRO paradigm. As mentioned above, participants also completed 2 other tasks in the scanner, and the order of the tasks was counterbalanced.

### MRI Data Preprocessing

Data were first converted from raw DICOM images to the BIDS specification format (Gorgolewski et al., 2016) using heudiconv (v0.5.1, https://github.com/nipy/heudiconv/releases/tag/v0.5.1). Data were then preprocessed using fMRIPrep (Esteban et al., 2019), a Nipype based tool (Gorgolewski et al., 2017, 2011). fMRIPrep performs anatomical and functional preprocessing basic steps (coregistration, normalization, unwarping, noise component extraction, segmentation, skullstripping, etc.). For each participant, the T1w (T1-weighted) volume was corrected for INU (intensity non-uniformity) using N4BiasFieldCorrection v2.1.0 (Tustison et al., 2010) and skull-stripped using antsBrainExtraction.sh v2.1.0 (using the OASIS template). Brain surfaces were reconstructed using recon-all from FreeSurfer v6.0.1 (Dale et al., 1999), and the brain mask estimated previously was refined with a custom variation of the method to reconcile ANTs-derived and FreeSurfer-derived segmentations of the cortical gray-matter of Mindboggle (Abraham et al., 2014a). Spatial normalization to the ICBM 152 Nonlinear Asymmetrical template version 2009c (Fonov et al., 2009) was performed through nonlinear registration with the antsRegistration tool of ANTs v2.1.0 (Avants et al., 2008), using brain-extracted versions of both T1w volume and template. Brain tissue segmentation of cerebrospinal fluid (CSF), white-matter (WM) and gray-matter (GM) was performed on the brain-extracted T1w using fast (Zhang et al., 2001), a part of FSL (FSL v5.0.9).

Functional data were motion corrected using mcflirt (FSL v5.0.9, Jenkinson et al., 2002). Distortion correction was performed using an implementation of the TOPUP technique (Andersson et al., 2003) using 3dQwarp (AFNI v16.2.07, Cox, 1996). This was followed by co-registration to the corresponding T1w using boundary-based registration (Greve and Fischl, 2009) with 9 degrees of freedom, using bbregister (FreeSurfer v6.0.1). Motion correcting transformations, field distortion correcting warp, BOLD-to-T1w transformation and T1w-to-template (MNI) warp were concatenated and applied in a single step using antsApplyTransforms (ANTs v2.1.0) using Lanczos interpolation. ICA-based Automatic Removal Of Motion Artifacts (AROMA) was used to create a variant of the data that is non-aggressively denoised (Pruim et al., 2015). AROMA uses a set of four robust, theoretically motivated temporal and spatial features to identify components related to head motion, and then uses linear regression to remove these components. Many internal operations of FMRIPREP use Nilearn (Abraham et al., 2014b), principally within the BOLD-processing workflow. For more details of the pipeline see https://fmriprep.readthedocs.io/en/latest/workflows.html.

Volume-based preprocessed data from FMRIPREP were then processed using CIFTIFY (v2.0.9, Dickie et al., 2019), a tool based on the Human Connectome Project (HCP) minimal preprocessing pipeline (Glasser et al., 2013). CIFTIFY allows for the processing of non-HCP datasets (i.e., data without T2w structural scans) using the Connectome Workbench (v1.3.2, https://github.com/Washington-University/workbench/releases/tag/v1.3.2). CIFTIFY used the MSMSulc method to align participants’ freesurfer-derived cortical surfaces (Robinson et al., 2018, 2014). Data was minimally smoothed with a 2mm FWHM Gaussian kernel, in line with the HCP minimally preprocessing pipeline. Subsequent data analysis was conducted with the HCP Pipelines (https://github.com/Washington-University/HCPpipelines/releases/tag/v4.0.0) using FSL (v6.0.1, Smith et al., 2004), FreeSurfer (v6.0.0, Dale et al., 1999), and the Connectome Workbench (v1.3.2, https://github.com/Washington-University/workbench/releases/tag/v1.3.2).

We converted whole-brain volumes to cortical surfaces and parcellated the group-level surfaces using the Multimodal Parcellation of Glasser and colleagues (2016) which consists of 180 regions in each hemisphere. This method has the added power of a region-of-interest analysis with high spatial sensitivity and whole-brain coverage. All analyses were carried out on data in the CIFTI format which stores data from cortical surfaces and subcortical volumes concurrently in a single file comprising a listed set of grayordinates. We conducted our analyses by parcellating each cortical surface into 180 regions using parcellation published by Glasser and colleagues (2016) MMP v1.0 cortical parcellation (2016).

This parcellation approach has several advantages: instead of correcting over ∼32k vertices, only 360 univariate analyses are performed, thus increasing sensitivity and statistical power; furthermore, because only minimal smoothing is applied (2 mm) there is limited blurring across regions from activated regions to adjacent, non-activated regions. Rather than restricting our analyses to *a priori* prefrontal cortical regions, we analyzed the whole MMP parcellation in order to have a hybrid region-of-interest/ whole-cortex analysis. A full-brain voxelwise timeseries analysis was conducted using the full CIFTI dense grayordinate data to investigate the contributions from subcortical regions and to compare to traditional volume-based analyses. These results are available on BALSA and in the Supplemental Results.

As we were trying to replicate Cole and colleagues (2010), we also conducted a confirmatory ROI analysis using parcels that corresponded to the clusters identified by Cole and colleagues They focused on a Right DLPFC ROI and a Left aPFC/RLPFC ROI, for which they reported Talairach coordinates of 29.6, 26.5, 35.3 and -22.3, 48.1, 18.5, respectively. For the Right DLPFC, the closest coordinate on the surface was 43.0, 27.4, 35.5, which was located at the border of MMP Areas R_8C and R_p-9-46v; we averaged Contrast Parameter Estimates (COPEs) from these two regions to create a Right DLPFC ROI. For the Left RLPFC, the closest coordinate on the surface was -34.3, 49.6, 18.9, which was located on the border of MMP Areas L_9-46d and L_a9-46v; we averaged COPEs from these two regions to create a Left RLPFC ROI. The COPE values for these 2 ROIs were entered into a Bayesian Repeated Measures ANOVA using JASP (0.11.1, 2019) with additional factors of group (HC, CHR), period (cue, trials), and condition (novel, practiced).

### fMRI Data Analysis

For the analysis of the preprocessed fMRI data, we modeled the task cue encoding period and the task performance period within the same model. We created 4 regressors of interest: novel cues, practiced cues, novel task performance, practiced task performance. Cues were modeled with a duration of 2.76s (i.e., the duration of presentation for all 3 cue rules) and trials were modeled with a duration of 1.84s (i.e., the duration of a single trial), and all regressors were convolved with the double-gamma hemodynamic response function with a temporal derivative. Errors were defined for task cues and task performance, with a task cue error categorized as a task cue followed by 2 or more incorrect trials. Eight contrasts were defined: Novel Cues > Practiced Cues, Practiced Cues > Novel Cues, Novel Cues Only, Practiced Cues Only, Novel Trials > Practiced Trials, Practiced Trials > Novel Trials, Novel Trials Only, Practiced Trials Only. First-level (within-run) modeling was carried out separately for each of the 6 runs; data were smoothed to a total of 4 mm FWHM, highpass filtered at 100s, and FILM prewhitening was used to account for temporal autocorrelation (Woolrich et al., 2001). A run was only included in higher-level analyses if it contained 3 or more novel or practiced blocks with 2 or more correct trials. The valid runs were averaged together in a between-runs fixed effects analysis. Within- and between-run analyses were carried out using the HCPP TaskfMRIAnalysisBatch pipeline.

Group-level statistics were performed using Permutation Analysis of Linear Models (PALM) (PALM v115a, Winkler et al., 2014), which is capable of computing univariate and multivariate non-parametric statistics using permutations and/or sign-flipping. Four group-level contrasts were defined: HC > CHR, CHR > HC, HC Mean, CHR Mean. In addition, a separate model was set-up to calculate the mean of all participants. To prepare the data for PALM, we concatenated the between-run participant outputs of each of the eight lower-level contrasts (Novel Cues > Practiced Cues, Practiced Cues > Novel Cues, Novel Cues Only, Practiced Cues Only, Novel Trials > Practiced Trials, Practiced Trials > Novel Trials Novel Trials Only, Practiced Trials Only). Each of the eight concatenated files was entered as a separate input and the results were corrected across the 8 within-subject map inputs and the 4 group-level contrasts to a Family-Wise Error Rate of .05. Separate PALM analyses were run for the left and right cortical surface. Results were saved as -log_10_(*p*), such that the minimum value considered was 1.60 (i.e., -log(.05/2)), to account for 2 tests. For the group comparison model, a mixture of 500 permutations and sign-flips were performed with tail approximation. For the analysis of the mean of all participants, 500 sign-flips were performed with tail approximation To further examine within and between group effects, we examined Cohen’s d maps saved by PALM (-saveglm option).

## Results

### Behavioral Results

We were interested in whether the HC and CHR participants learned at different rates during the practice session, as a group difference during training might account for how well the brain represented the practiced tasks. For the practice session, there was a large effect of block, with accuracy increasing over the course of training (*F*(4.59,201.98)=28.0, *p*<.001, η^2^_p_=0.39) and reaction time decreasing (*F*(3.16,138.94)=14.2, *p*<.001, η^2^_p_=0.24). While there was no interaction of block and group for accuracy (*F*(4.59,201.98)=0.42, *p*=.82, η^2^_p_=0.01), the HC group showed a larger decrease in reaction time compared to the CHR group (*F*(3.16,138.94)=3.03, *p*=.029, η^2^_p_=0.065).

During the scanning session, participants responded to novel tasks more slowly and less accurately compared to practiced tasks (Reaction time: *F*(45)=5.62, *p*<.001, Cohen’s d=0.83; Accuracy: Wilcoxon *W*=245, *p*<.004, Cohen’s d=-0.48). However, there was no interaction of task type and group (Reaction time: *F*(1,44)=0.04, *p*=.84, η^2^_p_=0.001; Accuracy: *F*(1,44)=0.65, *p*=.42, η^2^_p_=0.02). To examine whether there was further learning during the scanning session, we analyzed behavior over the course of the six scanner blocks. While participants became faster over the course of the scanning session (*F*(4.05, 174.36)=8.6, *p*<.001, η^2^_p_=0.167), there was no block by task type interaction (*F*(4.01, 172.56)=0.81 *p*=.52, η^2^_p_=0.018). Accuracy did not differ by block (*F*(4.01, 176.44)=1.47 *p*=.21, η^2^_p_=0.03). There were no effects with group for reaction time (all *F*’s < 0.81) or accuracy (all *F*’s < 0.48). Descriptive statistics are reported in Table 2 for reaction times and Table 3 for accuracy.

**Table 2.** Descriptive statistics for task performance, reaction times.

|  |  |  |  |  | 95% Confidence Interval |  |
| --- | --- | --- | --- | --- | --- | --- |
| Condition | Block | Group | Mean | SE | Lower | Upper |
| CHR | Novel | 1 | 1311 | 56 | 1200 | 1423 |
|  |  | 2 | 1276 | 56 | 1165 | 1387 |
|  |  | 3 | 1229 | 56 | 1118 | 1341 |
|  |  | 4 | 1215 | 56 | 1103 | 1326 |
|  |  | 5 | 1237 | 56 | 1125 | 1348 |
|  |  | 6 | 1205 | 56 | 1094 | 1317 |
|  | Practiced | 1 | 1255 | 56 | 1144 | 1367 |
|  |  | 2 | 1223 | 56 | 1112 | 1334 |
|  |  | 3 | 1196 | 56 | 1084 | 1307 |
|  |  | 4 | 1199 | 56 | 1087 | 1310 |
|  |  | 5 | 1196 | 56 | 1084 | 1307 |
|  |  | 6 | 1171 | 56 | 1059 | 1282 |
| HC | Novel | 1 | 1371 | 55 | 1261 | 1482 |
|  |  | 2 | 1305 | 55 | 1194 | 1416 |
|  |  | 3 | 1309 | 55 | 1198 | 1419 |
|  |  | 4 | 1294 | 55 | 1183 | 1405 |
|  |  | 5 | 1223 | 55 | 1113 | 1334 |
|  |  | 6 | 1217 | 55 | 1106 | 1327 |
|  | Practiced | 1 | 1322 | 55 | 1211 | 1433 |
|  |  | 2 | 1283 | 55 | 1173 | 1394 |
|  |  | 3 | 1249 | 55 | 1138 | 1359 |
|  |  | 4 | 1210 | 55 | 1100 | 1321 |
|  |  | 5 | 1201 | 55 | 1091 | 1312 |
|  |  | 6 | 1231 | 55 | 1121 | 1342 |

**Table 3.**
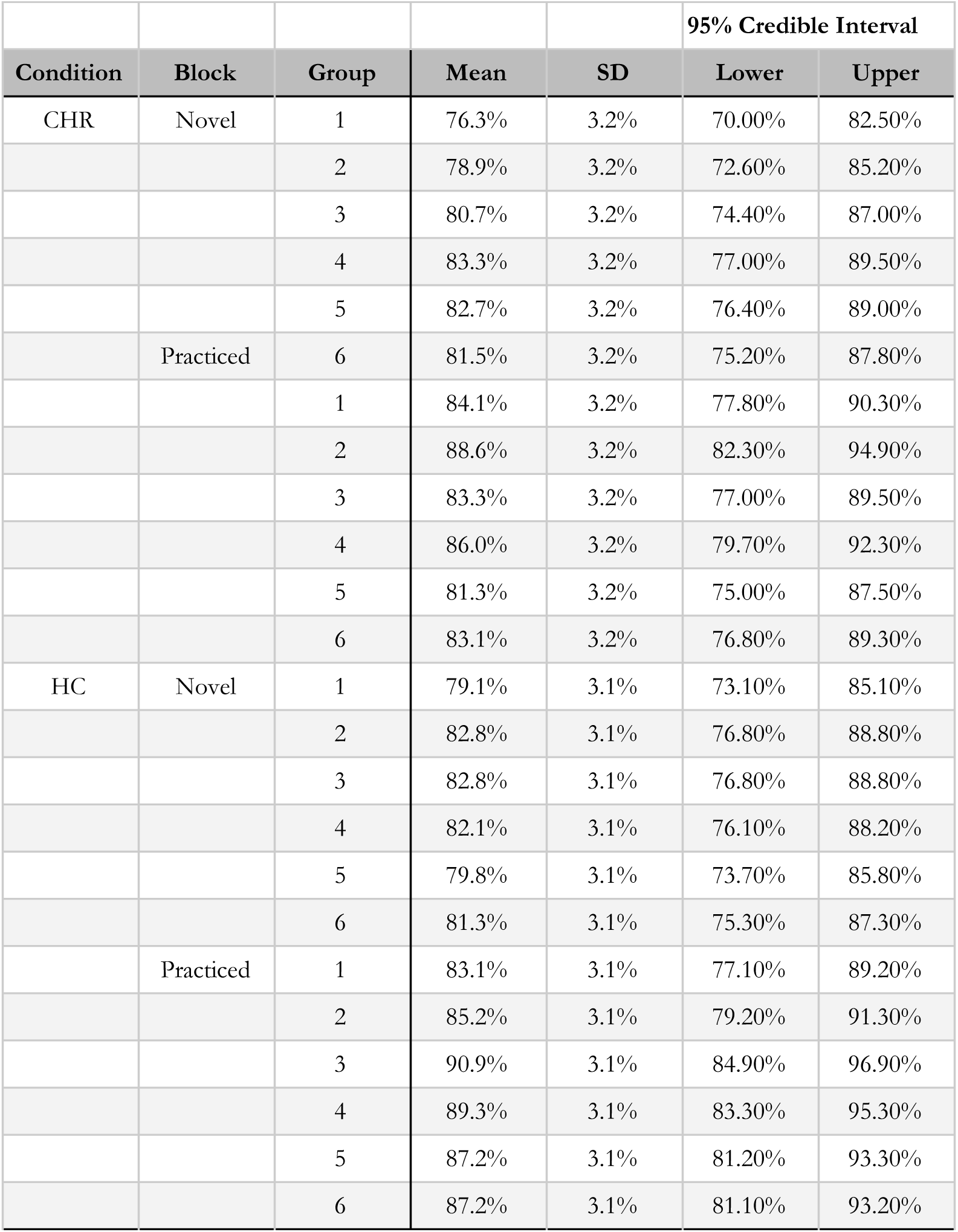
Descriptive statistics for task performance, accuracy.

We examined whether or not the groups differed in amount of head motion. We determined the average framewise displacement across each run, using the confounds output by fmriprep. The groups did not differ in terms of framewise displacement (CHR group: 0.098, HC group: 0.094; *F*(1,42,7) = 0.06, *p* = .81).

### Region-of-Interest Results

As noted in the Methods, the ROI analysis was performed using a Bayesian Repeated Measures ANOVA, with repeated factors of region (R_DLPFC, L_RLPFC), period (cue, trials), and condition (novel, practiced) and a between subject factor of group. When examining the model comparison, the best performing model compared to the Null model was the Region + Period model (BF_10_ = 2.16e+10). As shown in Table 4, the 3-way interaction of Region*Period*Condition, which would replicate the prefrontal dynamics reported by Cole et al. (2010), was not supported, with strong evidence in favor of accepting the null hypothesis (BF_incl_ = 0.067). There was, however, extreme evidence for the main effect of region (BF_incl_ = 3.26e+6 and period (BF_incl_>= 2.66e+11), as well as moderate evidence for a Period X Group interaction (BF_incl_ = 4.64). The CHR group showed a greater difference between the cue and trial period than the HC group. Of note, no effects involving Condition were supported (all BF’s < 0.78). As shown in Figure 2, there were no group differences in either region. The full model comparison is reported in Supplemental Table 1, the full analysis of effects is reported in Supplemental Table 2, and the full descriptive statistics are reported in Supplemental Table 3. Thus, the ROI analysis did not replicate the finding of prefrontal dynamics for novel and practiced tasks reported by Cole and colleagues (2010). Moveover, there was no support for the hypothesis that the prefrontal dynamics are blunted in CHR.

**Table 4.** Descriptive statistics from Region-of-Interest Analysis. Effects of region (L_DLPFC, R_RLPFC, R_aVLPFC), period (task cue encoding, task trial performance), condition (novel, practiced) by group (HC, CHR) on Contrast Parameter Estimates (COPE) were examined through a Bayesian Repeated Measures ANOVA.

|  |  |  |  |  |  |  | 95% Credible Interval |  |
| --- | --- | --- | --- | --- | --- | --- | --- | --- |
| Region | Period | Condition | Group | Mean | SD | N | Lower | Upper |
| L_RLPFC | Cue | Novel | CHR | 0.52 | 0.58 | 22 | 0.26 | 0.78 |
|  |  |  | HC | 0.47 | 0.81 | 24 | 0.13 | 0.81 |
|  |  | Practiced | CHR | 0.38 | 0.55 | 22 | 0.14 | 0.62 |
|  |  |  | HC | 0.56 | 0.67 | 24 | 0.28 | 0.84 |
|  | Trials | Novel | CHR | 0.89 | 0.68 | 22 | 0.59 | 1.19 |
|  |  |  | HC | 0.68 | 0.56 | 24 | 0.45 | 0.92 |
|  |  | Practiced | CHR | 0.75 | 0.68 | 22 | 0.45 | 1.05 |
|  |  |  | HC | 0.46 | 0.51 | 24 | 0.25 | 0.68 |
| R_DLPFC | Cue | Novel | CHR | 0.73 | 0.69 | 22 | 0.43 | 1.04 |
|  |  |  | HC | 0.76 | 0.87 | 24 | 0.40 | 1.13 |
|  |  | Practiced | CHR | 0.61 | 0.54 | 22 | 0.37 | 0.85 |
|  |  |  | HC | 0.78 | 0.67 | 24 | 0.50 | 1.07 |
|  | Trials | Novel | CHR | 1.16 | 0.63 | 22 | 0.88 | 1.44 |
|  |  |  | HC | 1.23 | 0.79 | 24 | 0.89 | 1.56 |
|  |  | Practiced | CHR | 1.00 | 0.62 | 22 | 0.73 | 1.28 |
|  |  |  | HC | 1.01 | 0.72 | 24 | 0.71 | 1.32 |

**Figure 2.**
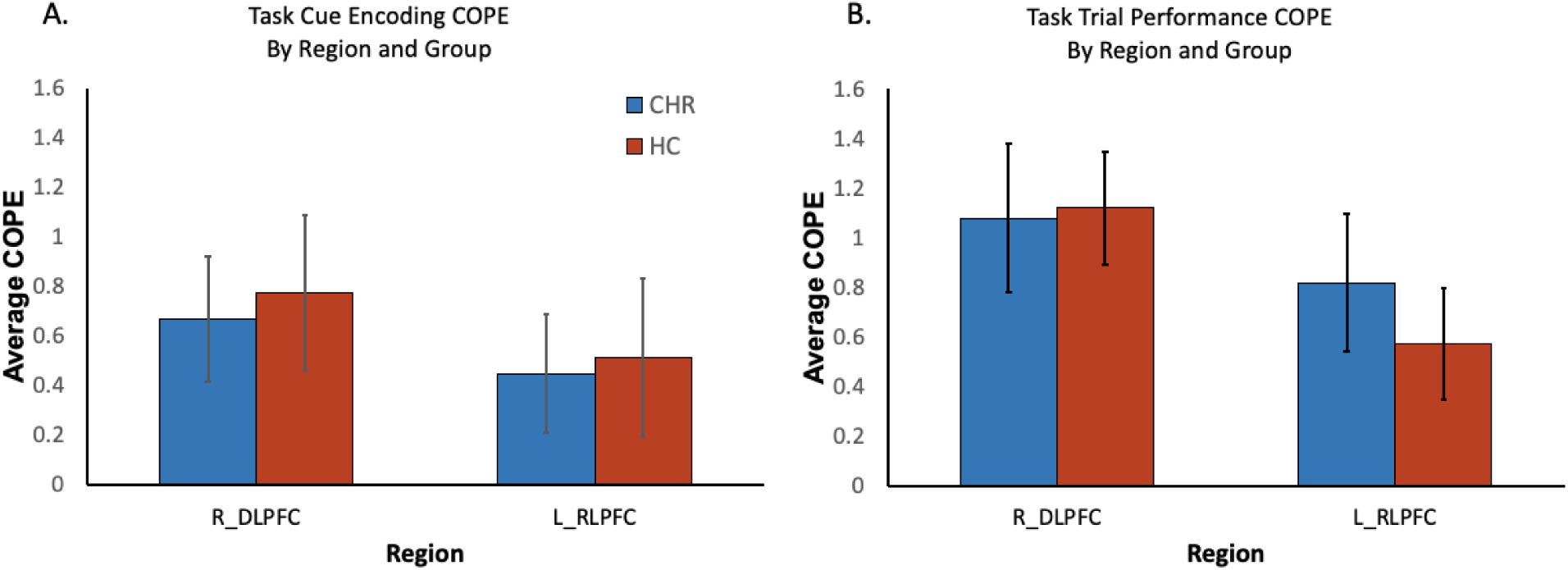
Region of interest analysis for task cue encoding (A) and task trial performance (B) by region and group. Contrast Parameter Estimates (COPE) were averaged across all the vertices within an ROI. Error bars represent 95% Credible Intervals.

### Within-Group Imaging Results

We first examined brain activation during task cue encoding and task performance with parcel-wise permutation statistical testing. As no parcels showed any significant differences in the contrast between novel and practiced tasks, we examined which parcels were significantly active in each condition alone (see Figure 3). While Cole and colleagues (2010) found that novel task encoding activated the Right DLPFC and practiced task encoding activated the Left RLPFC, here we found overlapping activation of Left mid-DLPFC (MMP Areas L_p9/46v, L_IFSp) and posterior DLPFC (MMP Areas L_IFJa, L_IFJp). In Healthy Controls, the Left and Right RLPFC (MMP Areas L-9-46d, R_9-46d, R_46) and Right DLPFC (MMP Areas R_p9-46v, R_IFSp) were activated during practiced task encoding. alone, suggesting that both the DLPFC and RLPFC may still be required for practiced task encoding. Cole and colleagues (2010) found that the frontal dynamics observed during task cue encoding was reversed during task trial performance, with activation of the Left RLPFC during novel tasks and Right DLPFC during practiced tasks. Once again, we failed to replicate this pattern, with no activation of the RLPFC for either condition, and overlapping activation of Left and Right mid-DLPFC (MMP Area L_p9-46v, R_p9-46v), as well as Left middle/posterior VLPFC & DLPFC (MMP Areas L_IFSp, L_IFJa, L_IFJp). In the Clinical High Risk group, these patterns of results were largely the same, but with no RLPFC for either condition during either task period. In sum, we found no significant differences for novel or practiced tasks in either group during task cue encoding and task trial performance, and failed to replicate the prefrontal dynamics observed by Cole and colleagues.

**Figure 3.**
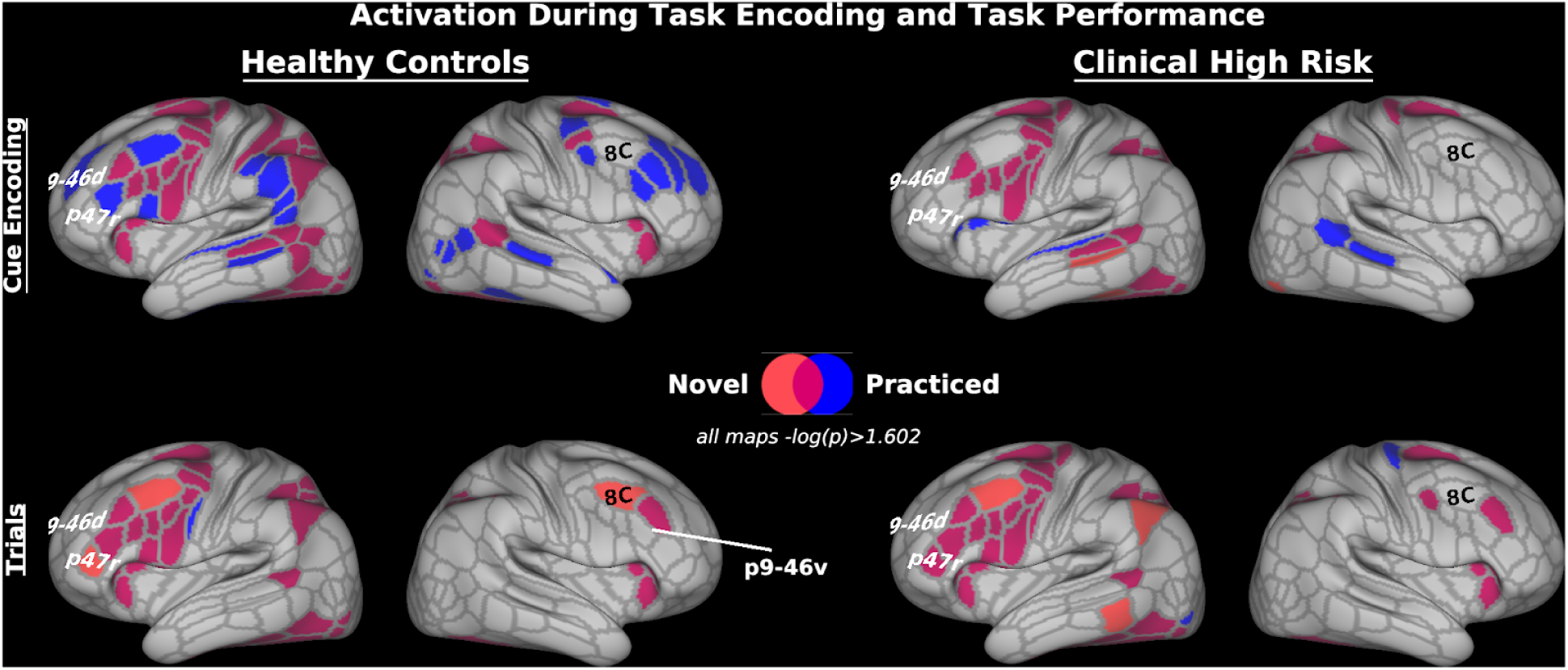
Activation during novel and practiced tasks at task cue encoding and task performance. Results are corrected at the parcel level with permutation testing to a threshold of -log(p) > 1.602 (equivalent to p<.05 with Bonferroni correction for 2 tests: Left Cortical Surface and Right Cortical Surface). Outlines depict the boundaries of the Glasser et al. Multi-modal Parcellation [91], and labels come from that parcellation. Novel trial activation is shown in red, practiced task activation is shown in blue, and overlapping activation is shown in purple. Outlines depict the boundaries of the Glasser et al. (2016) Multi-modal Parcellation, and labels come from that parcellation.

Since there were no significant results in the contrasts of novel and practiced trials, we examined the Cohen’s d effect size maps for the contrasts of novel and practiced task trials in each group (see Figure 4). These would indicate whether or not the design simply did not have enough power to identify significant effects. Partially supporting Cole and colleagues (2013, 2010), in the HC group, there was a small-to-moderate sized effect for the contrast of practiced > novel in the RLPFC (MMP Areas L_9-46d, L_a10p, L_9a) during task cue encoding. For task trial performance, there were large effects for the novel > practiced contrast in the anterior VLPFC (MMP Areas L_a10p, L_a9-46v), with medium-sized effects extending posteriorly to the Inferior Frontal Junction (MMP Areas L_IFJa, L_IFJp). This dorsal-ventral split for task cue encoding and task performance, respectively, has not been demonstrated with this task before. In the right hemisphere this pattern was largely replicated.

**Figure 4.**
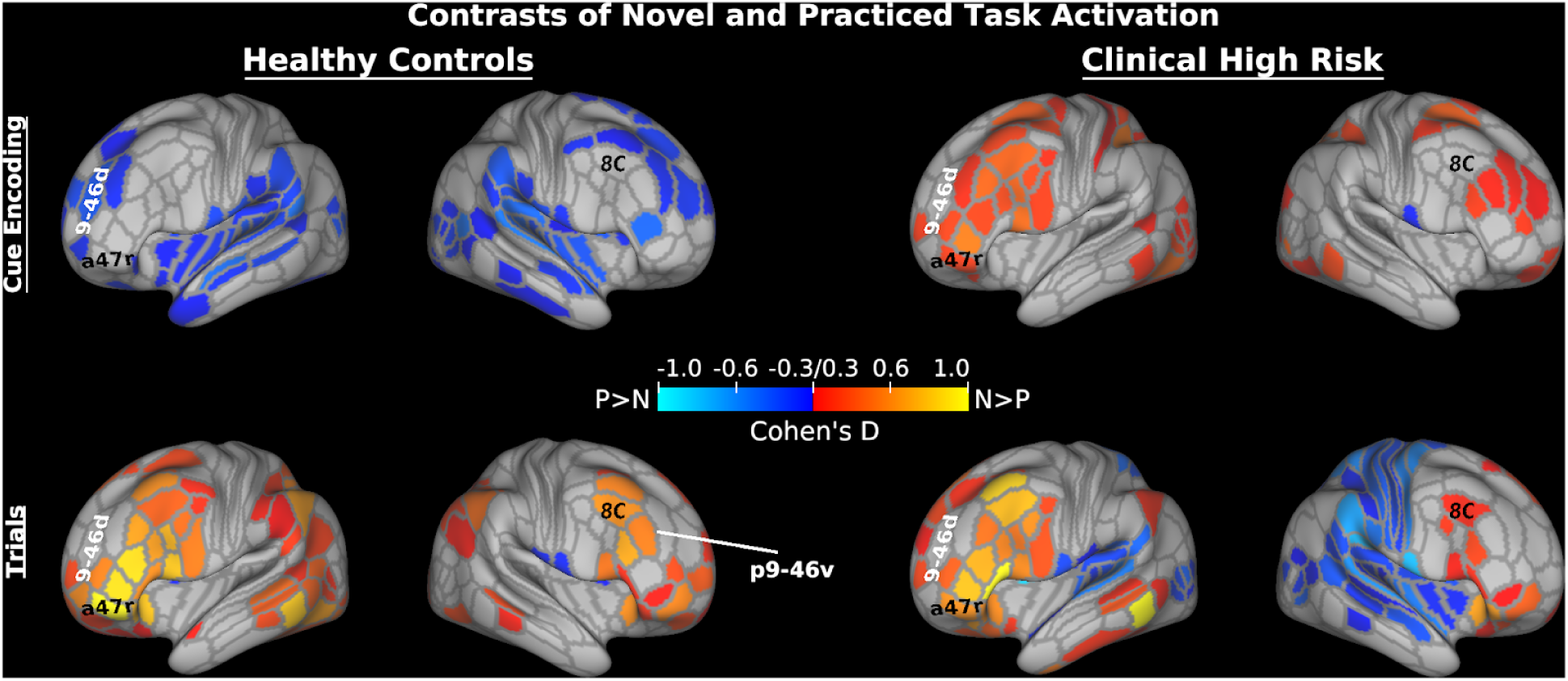
Effect size maps of contrasts of novel and practiced tasks at task cue encoding and task performance. The contrast of novel > practiced is shown in hot colors and the contrast of practiced > novel is shown in cold colors. Values shown are Cohen’s D with a minimum threshold of 0.3. Outlines depict the boundaries of the Glasser et al. (2016) Multi-modal Parcellation, and labels come from that parcellation.

Turning to the Clinical High Risk group, the task cue encoding period showed a much different pattern, with medium sized effects for the contrast of novel > practiced in the DLPFC and VLPFC. The CHR group largely resembled the HC group for task performance, albeit with weaker effects. Next, we directly compared the two groups.

### Between-group Imaging Results

As there were no statistically significant between-group contrasts, we examined the effect size maps for these contrasts. As shown in Figure 5, during task cue encoding, the prefrontal cortex showed small to large effects for the contrast of HC>CHR. The largest effects were observed in the DLPFC for practiced tasks. Moreover, controls showed large effects of activation across a number of key networks, including the fronto-parietal control network, motor regions, and higher-level visual regions, suggesting that controls were better at proactive control to prepare for a practiced task. The majority of regions with effects for the contrast of CHR>HC were observed during task performance; the regions with the largest effect were 9p and a9-46v. Notably, these regions were absent in the HC group, as shown in Figures 3 and 4.

**Figure 5.**
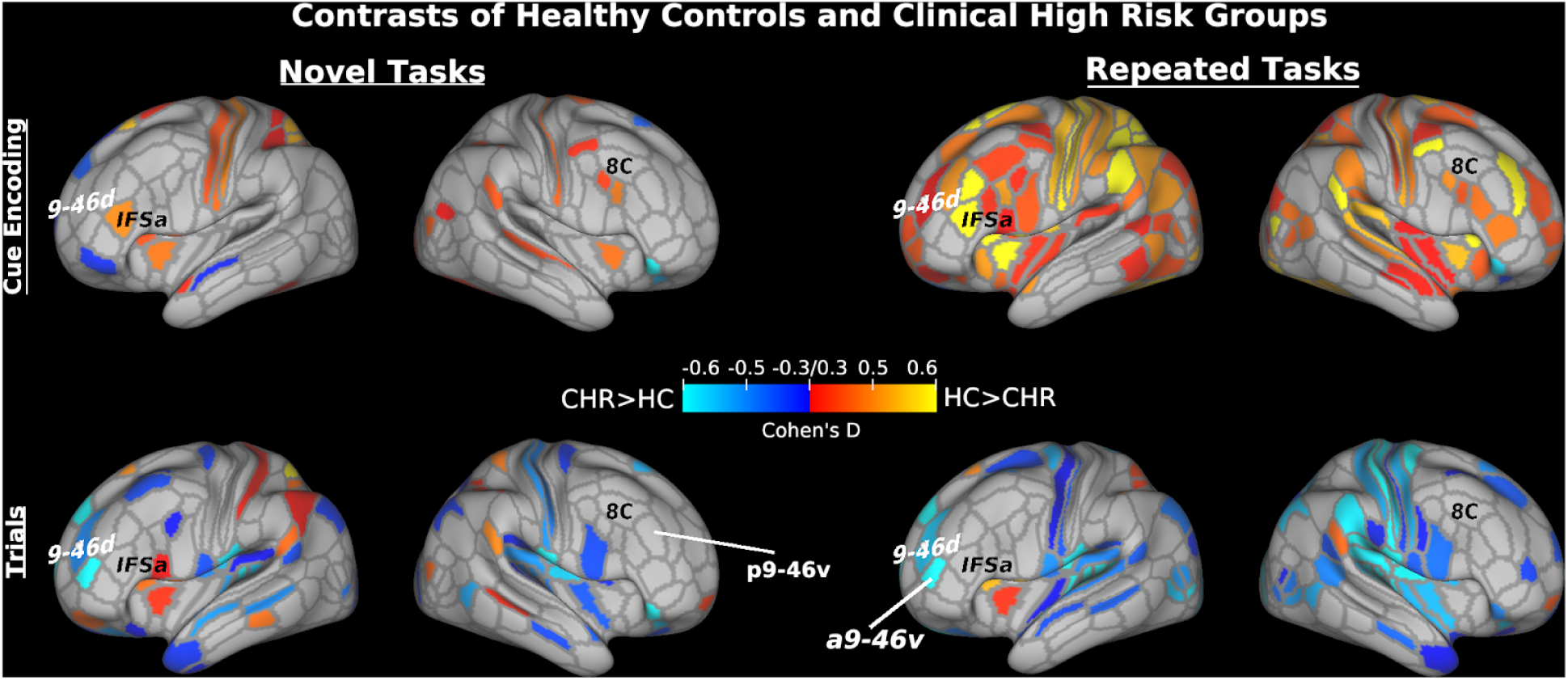
Effect size maps of contrasts of healthy control (HC) and clinical high risk (CHR) groups. The contrast of HC> CHR is shown in hot colors and the contrast of CHR > HC is shown in cold colors. Values shown are Cohen’s D with a minimum threshold of 0.3. Outlines depict the boundaries of the Glasser et al. (2016) Multi-modal Parcellation, and labels come from that parcellation.

### Across-Group Results

Lastly, we compared novel and practiced task activation for all 46 participants together. While there were no differences during task cue encoding, several parcels in the left anterior ventrolateral prefrontal cortex (aVLPFC) showed greater activation for novel vs. practed task trial performance (see Figure 6). Specifically, these parcels included rostral orbital frontal cortex (Areas L_a47r and L_p47r), anterior Inferior Frontal Sulcus (Area L_IFSa), and Area L_45. Notably, the RLPFC parcels (i.e., L_9-46d, L_a9-46v) did not show a significant difference, although these regions were significantly active during novel task trial performance but not practiced (see Supplemental Figure 10). These aVLPFC regions are functionally connected with anterior temporal cortex and anterior dorsomedial prefrontal cortex (Neubert et al., 2014), and have been suggested to play a role in semantic processing (Amunts et al., 2010), suggesting that these regions were involved in semantic processing of the task rules during task performance.

**Figure 6.**
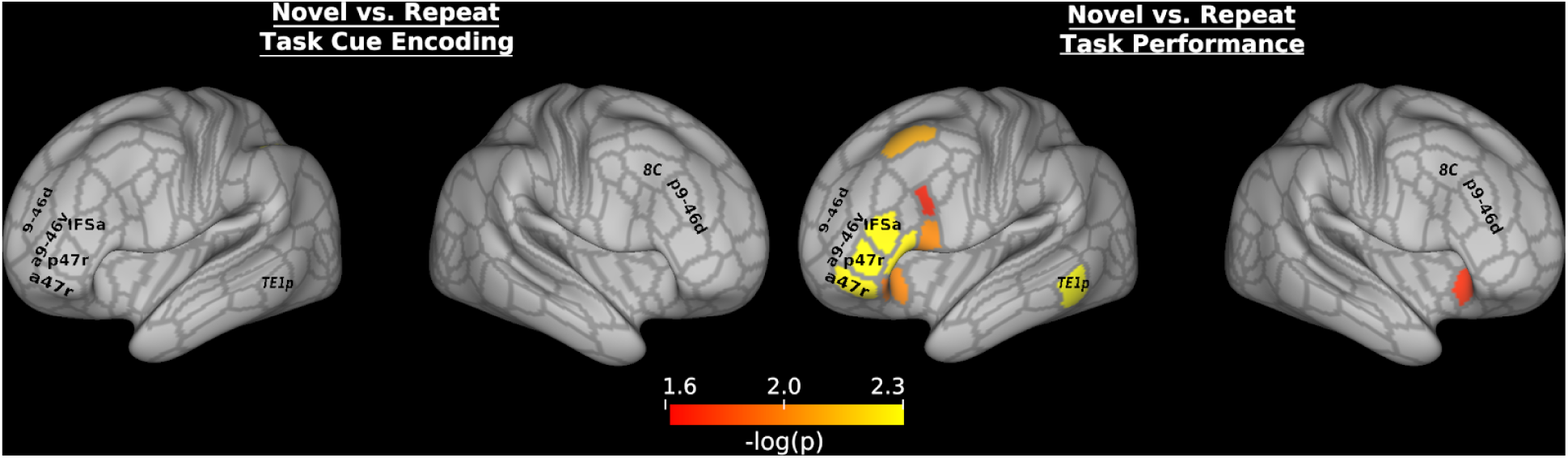
Contrasts of novel vs. practiced tasks during task cue encoding and task performance across both groups (Healthy Controls and Clinical High Risk). Results are corrected at the parcel level with permutation testing to a threshold of -log(p) > 1.602 (equivalent to p<.05 with Bonferroni correction for 2 tests: Left Cortical Surface and Right Cortical Surface.). Outlines depict the boundaries of the Glasser et al. (2016) Multi-modal Parcellation, and labels come from that parcellation.

## Discussion

In the current study we investigated rapid instructed task learning (RITL) in a group of adolescents at clinical high risk (CHR) for psychosis and a group of healthy control (HC) participants. Participants were required to quickly encode a set of rules into a goal set and then maintain this goal set to perform a series of trials (Cole et al., 2010). While previous studies have demonstrated that a disability in representing goal information is central to schizophrenia (Barch and Ceaser, 2012), it has been unclear if this deficit begins before or after the onset of schizophrenia.

Moreover, previous fMRI studies of goal maintenance in psychosis have focused on well learned tasks while in our day to day lives we may have to adapt previously acquired rules to new tasks and contexts. While no statistically significant group differences were identified, we identified moderate-to-large sized effects suggesting that CHR participants show poor task rule encoding, and rely on less effective reactive control mechanisms to apply task rules during task performance. Overall, this is a novel investigation of the course of goal maintenance deficits in psychosis and the first study of rapid task learning in psychosis.

### Rapid Instructed Task Learning in Controls

In healthy controls, we aimed to replicate previous findings by Cole and colleagues (2016, 2010) demonstrating a reversal of prefrontal brain dynamics for practiced and novel task encoding and performance. Cole and colleagues found that novel task rules are first encoded by the DLPFC and then integrated into working memory by the RLPFC during task performance; these dynamics are reversed for practiced tasks, with rules being retrieved from long-term memory by the RLPFC and then activated by DLPFC working memory mechanisms for task performance. Using high-resolution, ultra-fast multiband fMRI sequences and cutting-edge analysis techniques developed by the Human Connectome Project, we largely failed to replicate this pattern in healthy controls, and identified some critical deviations.

In an ROI analysis, we failed to find evidence for a main effect of condition (Novel vs. Practiced), or critically, an interaction of region*period*condition. When we examined the thresholded statistical maps in the current study, we found no statistically significant differences in activation for the contrast of novel vs. practiced tasks. Compared to baseline, novel and practiced tasks both showed significant activation of the RLPFC and DLPFC during task cue encoding and DLPFC and aVLPFC activation during task performance. However, when we looked at the effect size maps (Cohen’s D), the HC group showed a small effect of greater RLPFC activation for practiced vs. novel task cue encoding.

If the novel rules were truly represented as distinct from practiced rules, we would expect to see greater activation of brain areas involved in switching rules or switching tasks (Cole et al., 2010). Switching of tasks or task sets has been suggested to rely on regions of the lateral prefrontal cortex (primarily DLPFC and IFJ) as well as the intraparietal sulcus (IPS) (Kim et al., 2012). Rule switching, however, has been suggested to rely on medial rather than lateral prefrontal cortex (Crone et al., 2005). MVPA decoding work has shown that specific task rules are first encoded in the IPS, though this is likely to be true primarily for practiced rules (Bode and Haynes, 2009). Nevertheless, in the current study, there was little evidence to suggest a difference in how the novel and practiced rules were encoded.

The task performance data deviated further from the findings of Cole and colleagues. As with the task cue encoding results, we found no statistically significant differences between novel and practiced task performance. While the effect size maps showed small-to-medium sized effects for the contrast of Novel>Practiced, the effect was found in the anterior VLPFC. This region has been associated with general retrieval of rules from long-term memory (Cole et al., 2010; Donohue et al., 2005) as well as the need to control such retrieval mechanisms (Badre and Wagner, 2004). Supporting this role for the anterior VLPFC, we found the same effect in the middle temporal cortex. While we predicted that such retrieval mechanisms would be involved during task cue encoding, particularly for practiced tasks, it was surprising to see possible retrieval during task performance. One possibility is that participants retrieved instances from practiced task trials in order to determine how to apply the novel rule. For instance, participants may have practiced judging the sweetness of the stimuli or applying the logic rule of both stimuli requiring the same semantic label, and drew upon those instances to apply a novel task set. In support of this possibility, behavioral responses to novel tasks were slower and less accurate compared to practiced tasks, allowing for such a late, reactive control process to occur. As noted elsewhere, the psychological need for short cue-to-target intervals in typical task switching designs has made it difficult to separate preparatory activity locked to the cue and reactive activity locked to the target presentation (Ruge et al., 2013).

Further evidence for a role of the aVLPFC in representing task rules comes from the across-group analysis. Across both groups, the aVLPFC was significantly more active during novel vs. practiced task performance. This suggests that the novel tasks were performed by retrieving components of practiced task rules in order to apply the novel combination of rules on the current trials. As we used a slow-event related design with an average interval between the onset of the last cue stimulus and the first trial of 4.6s (and we analyzed all 3 trials, not just the 1st trial), it seems unlikely that the involvement of the aVLPFC on novel tasks arose during the cue period. This suggests that any additional processing during novel task performance compared to practiced task performance was not due to preparatory activity.

### Task Learning Deficits in Psychosis Risk

Qualitatively, the HC and CHR groups appeared most different during task cue encoding. As shown in Figure 4, the HC group showed a stronger effect for the contrast practiced > novel tasks, but the CHR group showed a stronger effect for the contrast novel > practiced tasks. However, when directly comparing the groups, there were little group contrast effects for novel task encoding, but there were large group effects during practiced task encoding for the contrast HC > CHR, albeit with no statistically significant group effects. The large effect size of the group difference across the DLPFC/VLPFC suggests that CHR participants use different strategies for retrieval of learned task rules (Badre et al., 2005; Kostopoulos and Petrides, 2003). This is despite the lack of a difference in performance during the practice session, so it is unlikely to reflect deficient learning of the practiced tasks.

During both novel and practiced task trial performance, we found moderate-to-large effects for the CHR > HC contrast in the RLPFC (namely areas 9-46d and a9-46v). These regions, in particular a9-46v, are thought to coordinate control processes (Badre, 2008; Badre and Nee, 2017) or more generally activate during increased cognitive demand (Assem et al., 2019; Crittenden and Duncan, 2012). The latter hypothesis in particular suggests that the CHR group needs additional resources to perform the tasks, perhaps due to worse preparation.

These findings are in line with previous studies showing poor proactive control and goal maintenance in schizophrenia in conjunction with deficits in prefrontal functioning (Barch and Ceaser, 2012; Poppe et al., 2016; Sheffield et al., 2014). Although less prevalent in the group comparison, the CHR group appeared to rely on reactive control mechanisms, in particular for novel task performance. Lesh and colleagues (2013) found that first-episode schizophrenia was associated with hypoactivation of the DLPFC during proactive control, but normal activation during reactive control. Reactive control has also been shown to be intact in schizophrenia in instances of motivated control in response to reward (Mann et al., 2013). Nevertheless, patients with schizophrenia show diminished activation of control regions at longer RTs, which has been suggested to reflect deficits of reactive control mechanisms needed to overcome lapses of proactive control (Fassbender et al., 2014). However, further research is needed to elucidate whether the activation of frontal and parietal control regions during task performance, as opposed to during cue encoding actually reflects reactive control.

Overall, there are some limited signs to suggest that the Clinical High Risk stage of psychosis is associated with alterations of goal maintenance and task set learning. It has been suggested that schizophrenia patients may use inefficient encoding and retrieval strategies compared to healthy controls (MacDonald et al., 2005). This may be due to a breakdown in networks that support the integration of long-term memory and working memory (Ragland et al., 2012). Although the CHR group may not have shown any reductions in activity during novel task encoding, the results suggest that the instructions were not encoded efficiently, forcing them to rely on retrieval mechanisms during task performance, rather than more efficient preparatory control. Future studies should use different tasks and larger samples to more definitively investigate whether those at clinical high risk have disrupted preparatory control and/or goal maintenance. With a moderate effect size (D=.5), in order to have 80% power to be able to detect a significant difference in mean activation between 2 groups, 51 participants would be needed in each group^1^.

### Limitations and Future Directions

Although our sample size was in line with recent studies of executive function in psychosis, the disparity between the effect size and the lack of significant group-level effects suggests that additional participants are needed. We used a recently developed parcellation approach which added power without losing spatial coverage. To this end, we were able to investigate the contributions of prefrontal subregions to executive functions. Although we adapted the task previously used by Cole and colleagues (2013, 2010), we used a different analysis approach. We did not employ a Finite Impulse Response model as used by Cole and colleagues. Cole and colleagues first identified regions within the prefrontal cortex that showed a condition (practiced vs. novel) X time interaction with a liberal cluster formation threshold and then performed ANOVAs on regions of interest to identify region X condition interactions during the encoding and task performance periods. When we extracted parameter estimates from similar ROIs, we did not find a region X condition X period interaction, reflecting a failure to directly replicate Cole and colleagues. Moreover, our effect size maps suggest that even with a different analysis strategy, neither the HC or CHR participants would show the pattern of results demonstrated by Cole and colleagues. Future studies should investigate whether psychosis risk participants show deficits in rule learning using different paradigms such as those developed by Dumontheil and colleagues (2011). Larger sample sizes will also enable researchers to possibly examine subgroups of psychosis risk; our sample consisted mostly of those included for Attenuated Positive Symptom Prodromal Syndrome, with only 2 participants who also met criteria for Schizotypal Personality Disorder, and no participants included for Genetic Risk and Deterioration Prodromal Syndrome, so we could not examine whether specific subgroups show worse cognitive functioning. Doing so will be critical for understanding the etiology of psychosis.

## Data Availability

https://openneuro.org/datasets/ds001371

https://balsa.wustl.edu/Klv19

## Acknowledgements

We would like to thank three anonymous reviewers for their thoughtful criticisms and suggestions on a previous version. We are grateful to Tina Gupta for assistance with participant recruitment and to Derek Dean and all of the research assistants at the CU ADAPT Program for their help with running participants. E-Prime file for the paradigm were graciously provided by Michael W. Cole. This work was funded by the National Institute of Mental Health R01MH094650 to V.A.M.. J.M.O. was supported by F32DA034412 and by start-up funds from Texas A&M University. J.A.B. was supported by NIH F32MH102898, a Brain & Behavior Research Foundation NARSAD Young Investigator Award as the Donald & Janet Boardman Family Investigator, and by start-up funds from Texas A&M University.

## Supplemental Figures

**Supplemental Figure 1.**
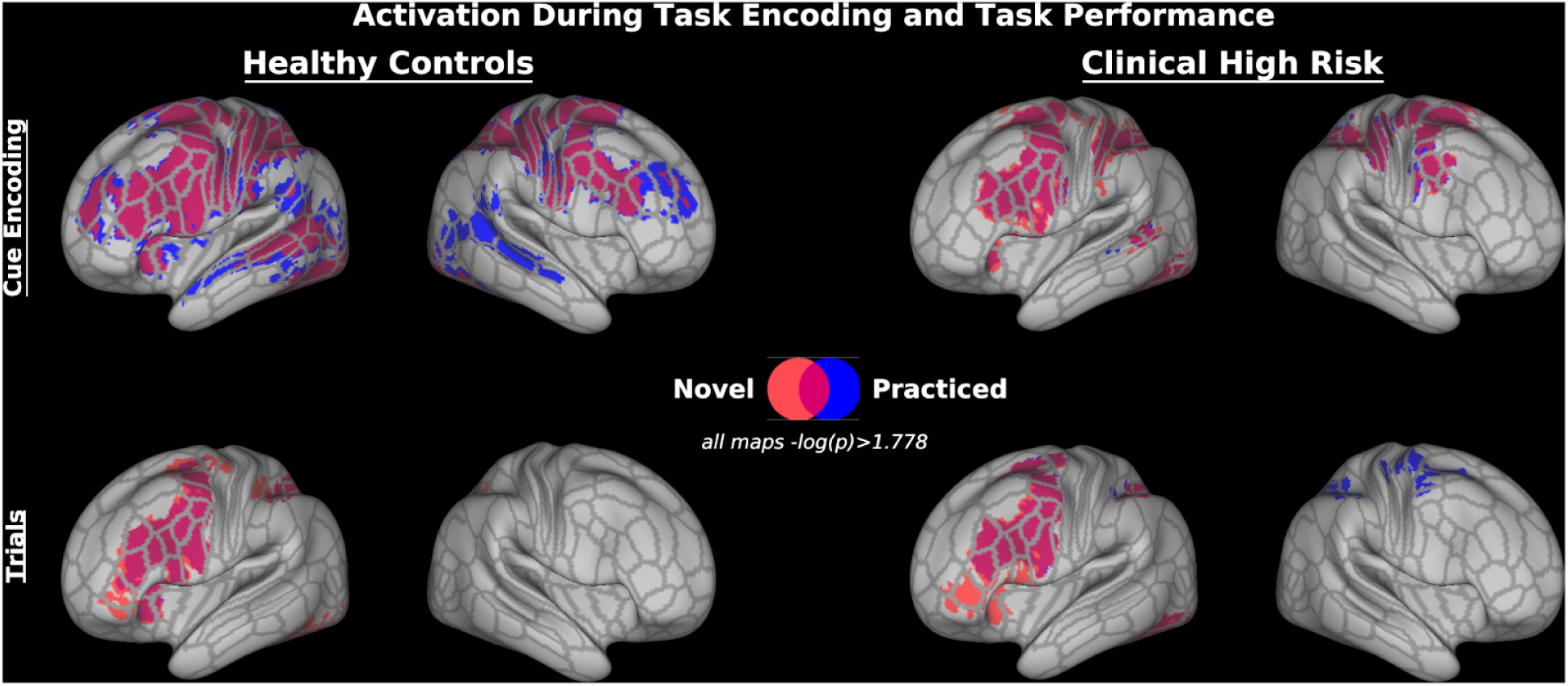
Dense grayordinate surface activation during novel and practiced tasks at task cue encoding and task performance. Results are corrected at the cluster level with threshold-free cluster enhancement (TFCE) and permutation testing to a threshold of -log(p) > 1.778 (equivalent to p<.05 with Bonferroni correction for 3 tests: Left Cortical Surface and Right Cortical Surface, Subcortical Volume). Outlines depict the boundaries of the Glasser et al. (2016) Multi-modal Parcellation. Novel trial activation is shown in red, practiced task activation is shown in blue, and overlapping activation is shown in purple.

**Supplemental Figure 2.**
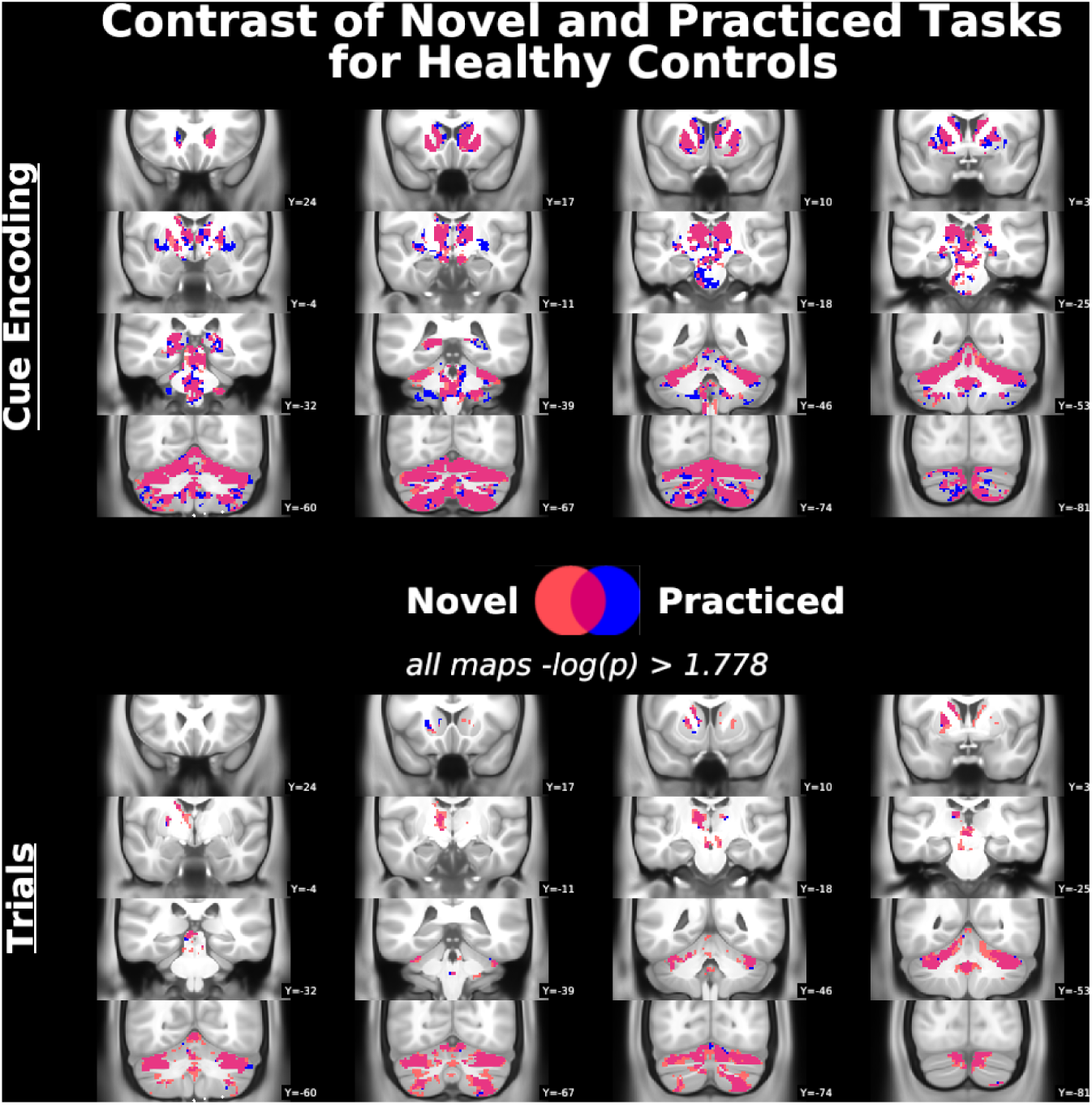
Full subcortical volume results of the analysis of activation during novel and practiced tasks at task cue encoding and task performance in the Healthy Control Group. Results were corrected using Threshold-Free Cluster Enhancement with permutation testing to a threshold of -log(p) > 1.778, which is equivalent to p<.05 with Bonferroni correction for 3 tests: Left Cortical Surface, Right Cortical Surface, subcortical volume. Novel trial activation is shown in red, practiced task activation is shown in blue, and overlapping activation is shown in purple.

**Supplemental Figure 3.**
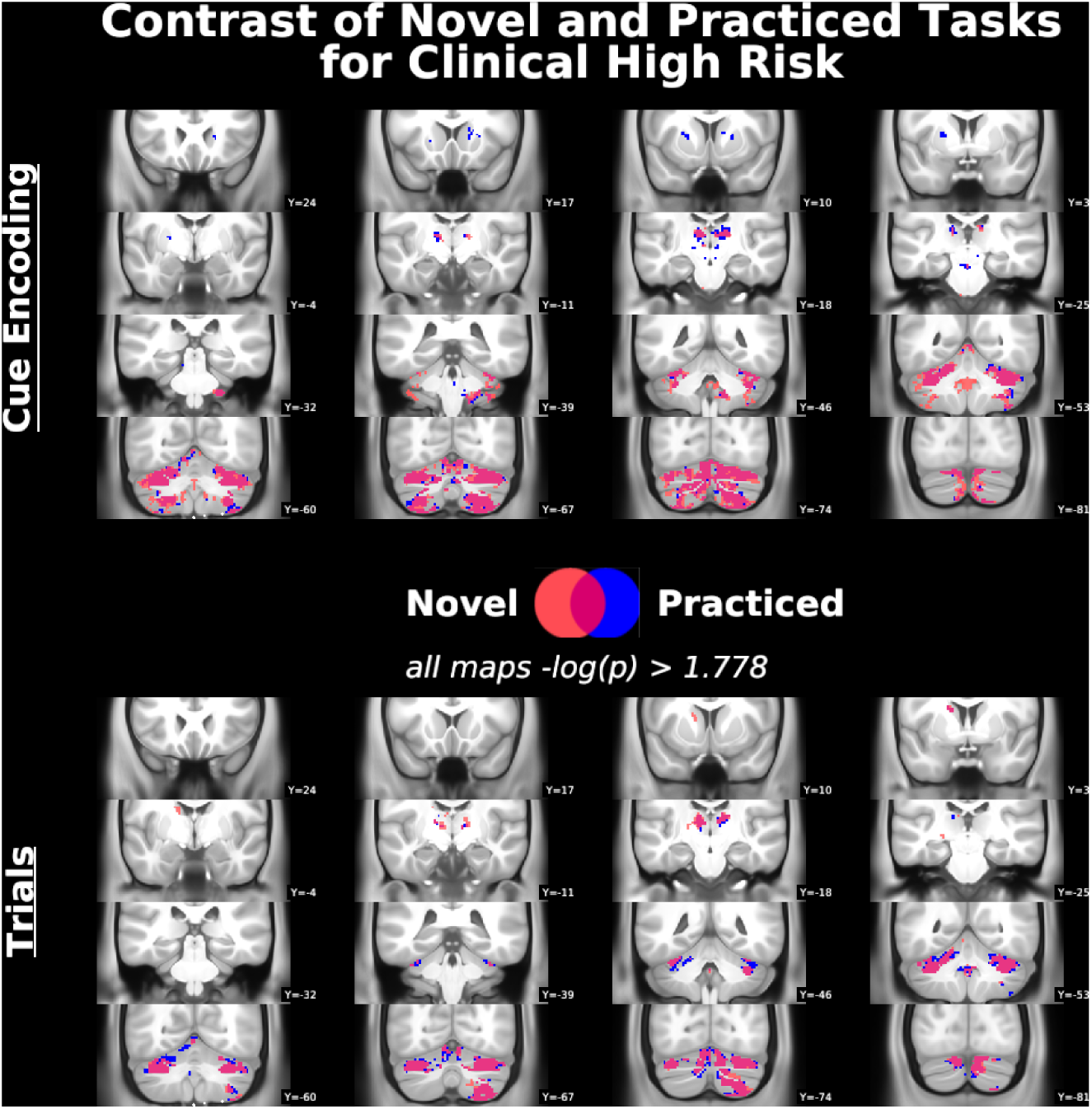
Full subcortical volume results of the analysis of activation during novel and practiced tasks at task cue encoding and task performance in the Clinical High Risk Group. Results were corrected using Threshold-Free Cluster Enhancement with permutation testing to a threshold of -log(p) > 1.778, which is equivalent to p<.05 with Bonferroni correction for 3 tests: Left Cortical Surface, Right Cortical Surface, subcortical volume. Novel trial activation is shown in red, practiced task activation is shown in blue, and overlapping activation is shown in purple.

**Supplemental Figure 4.**
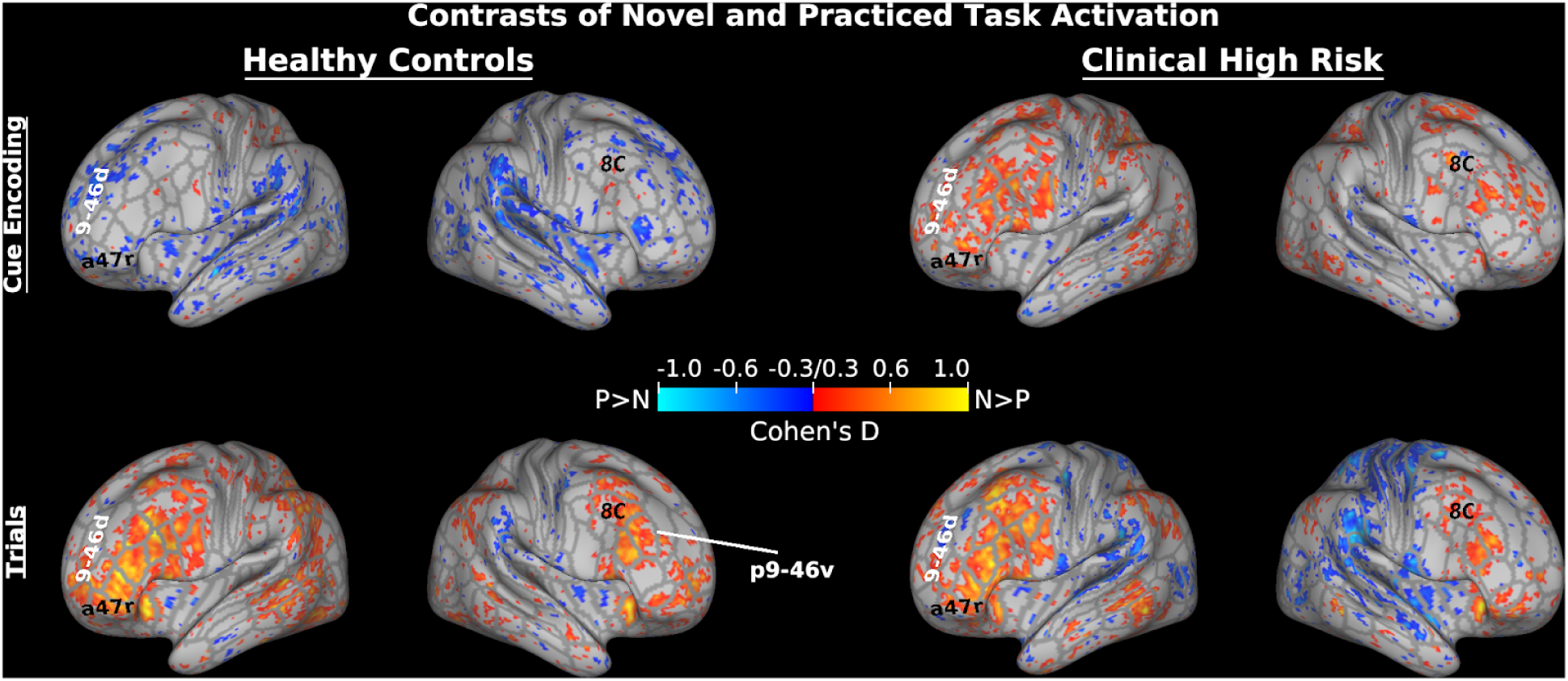
Dense grayordinate effect size maps of contrasts of novel and practiced tasks at task cue encoding and task performance. The contrast of novel > practiced is shown in hot colors and the contrast of practiced > novel is shown in cold colors. Values shown are Cohen’s D with a minimum threshold of 0.3. Outlines depict the boundaries of the Glasser et al. (2016) Multi-modal Parcellation, and labels come from that parcellation.

**Supplemental Figure 5.**
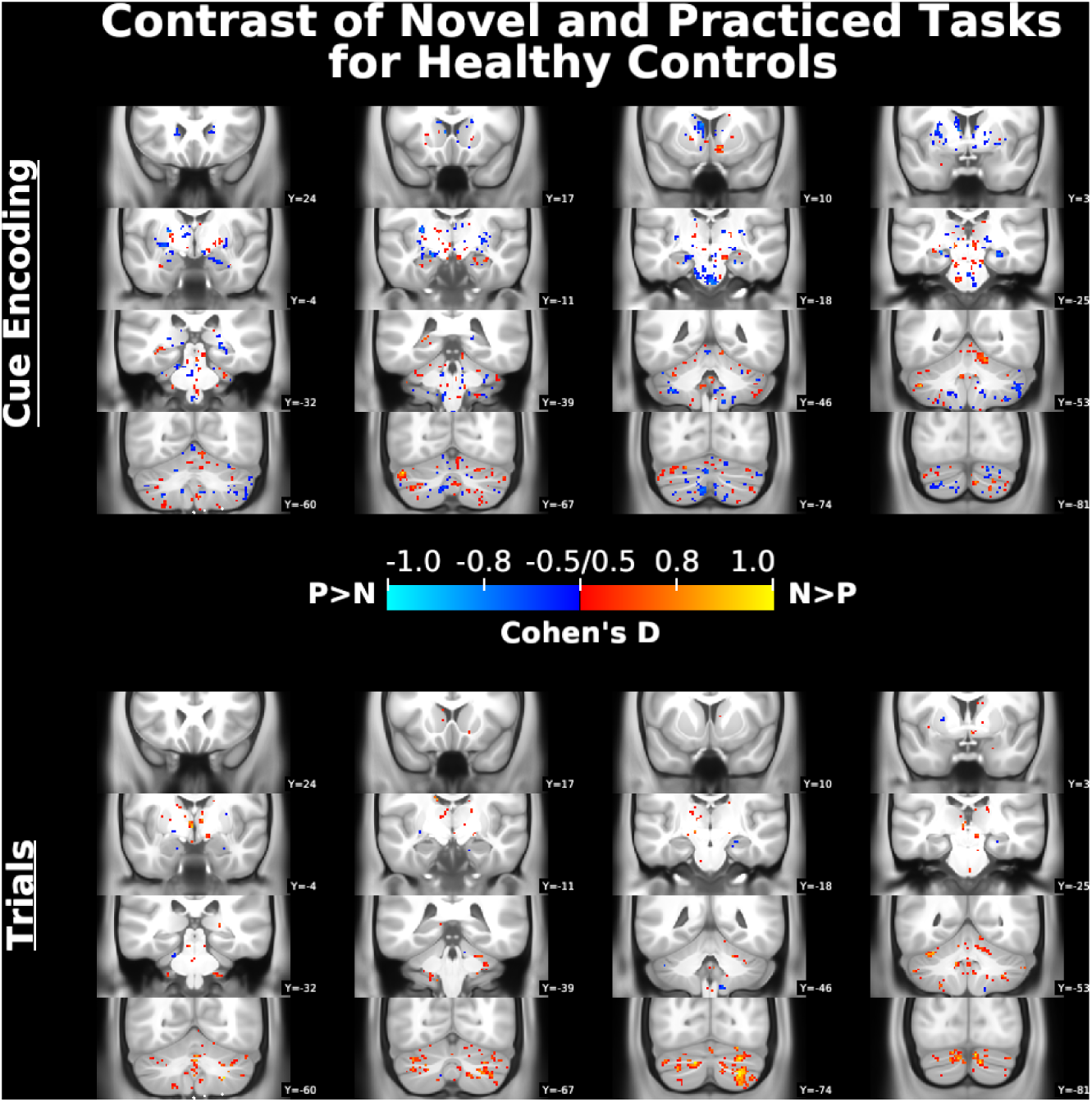
Subcortical effect size maps of contrasts of novel and practiced tasks at task cue encoding and task performance in the Healthy Controls Group. The contrast of novel > practiced is shown in hot colors and the contrast of practiced > novel is shown in cold colors. Values shown are Cohen’s D with a minimum threshold of 0.3.

**Supplemental Figure 6.**
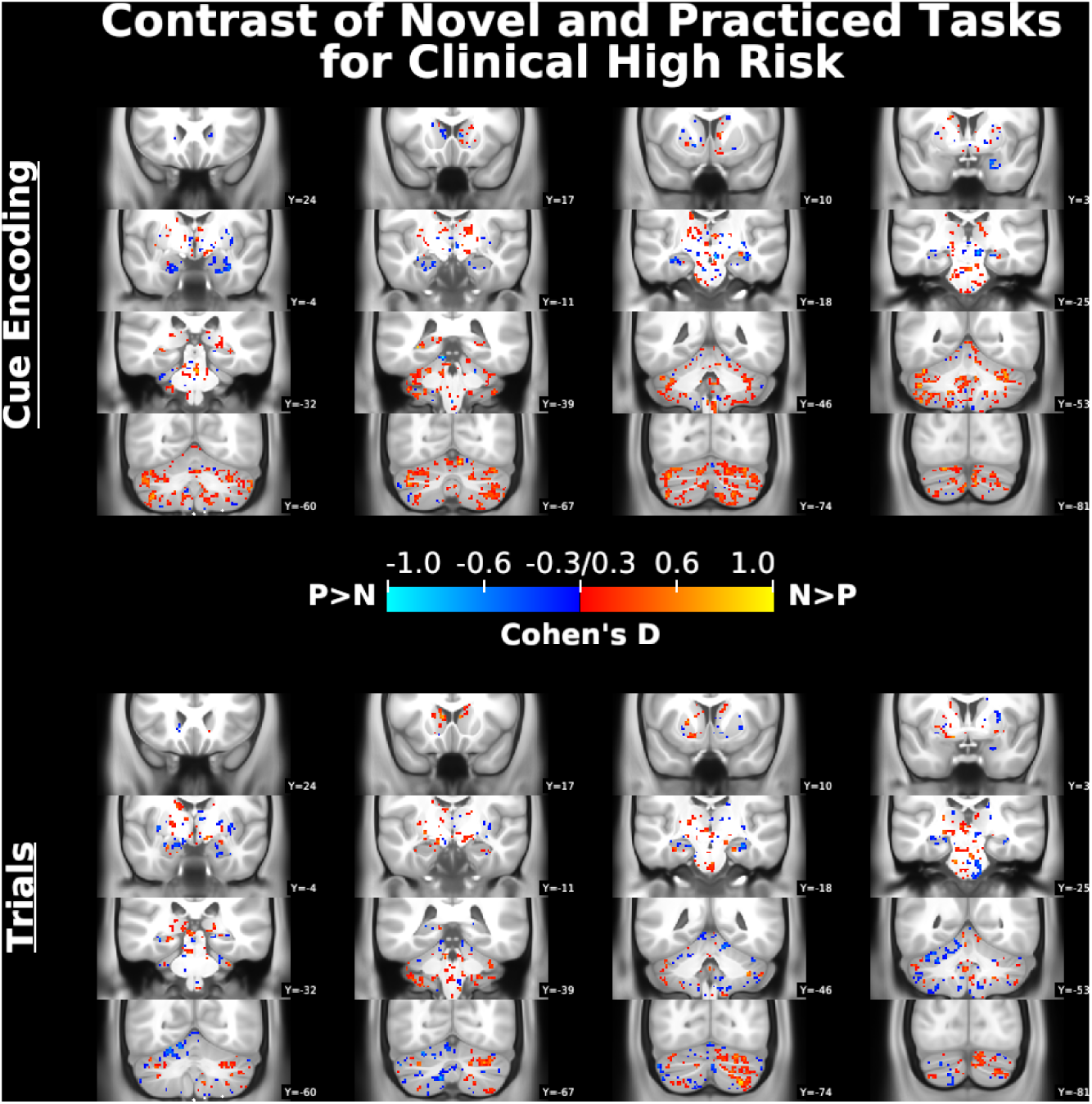
Subcortical effect size maps of contrasts of novel and practiced tasks at task cue encoding and task performance in the Clinical High Risk Group. The contrast of novel > practiced is shown in hot colors and the contrast of practiced > novel is shown in cold colors. Values shown are Cohen’s D with a minimum threshold of 0.3.

**Supplemental Figure 7.**
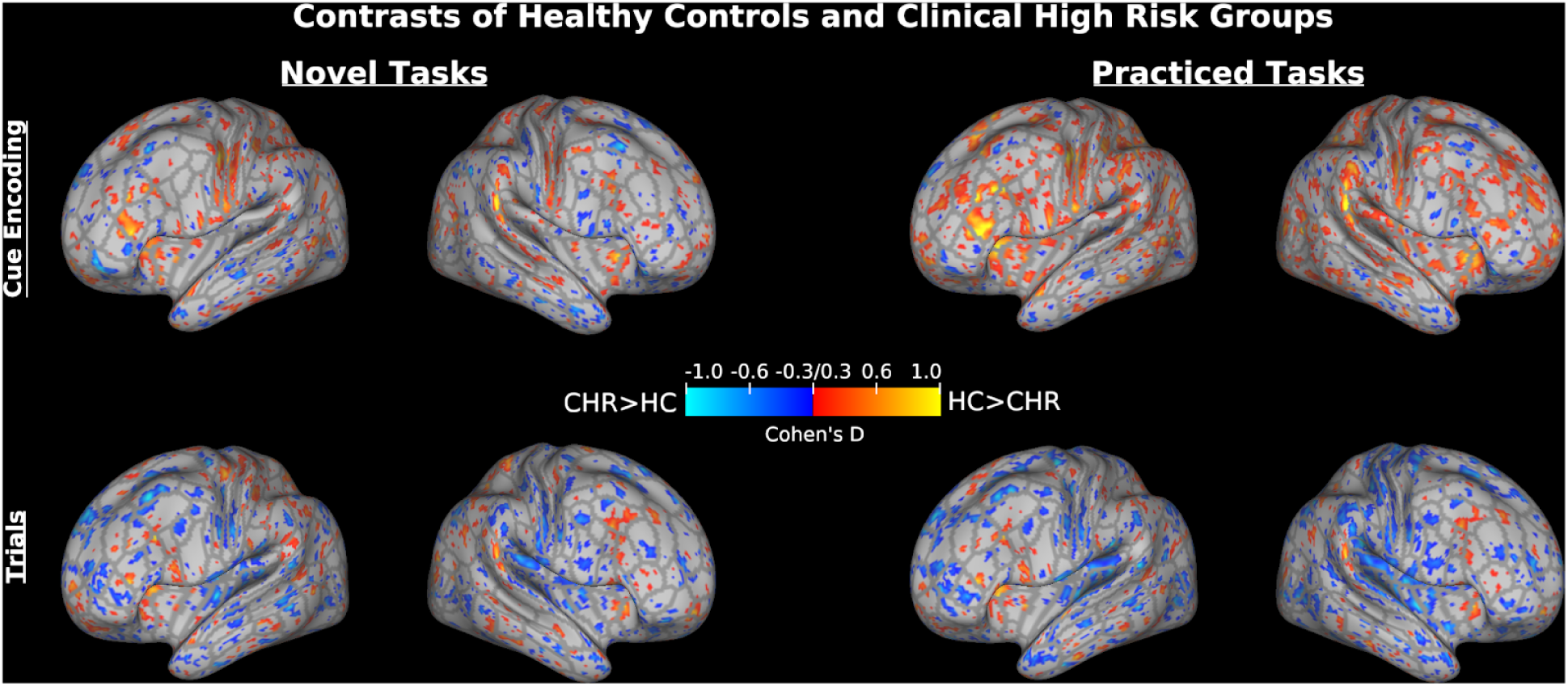
Dense grayordinate effect size maps of contrasts of the Healthy Control (HC) group and the Clinical High Risk (CHR) group for novel and practiced tasks at task cue encoding and task performance. The contrast of HC > CHR is shown in hot colors and the contrast of CHR > HC is shown in cold colors. Values shown are Cohen’s D with a minimum threshold of 0.3. Outlines depict the boundaries of the Glasser et al. (2016) Multi-modal Parcellation.

**Supplemental Figure 8.**
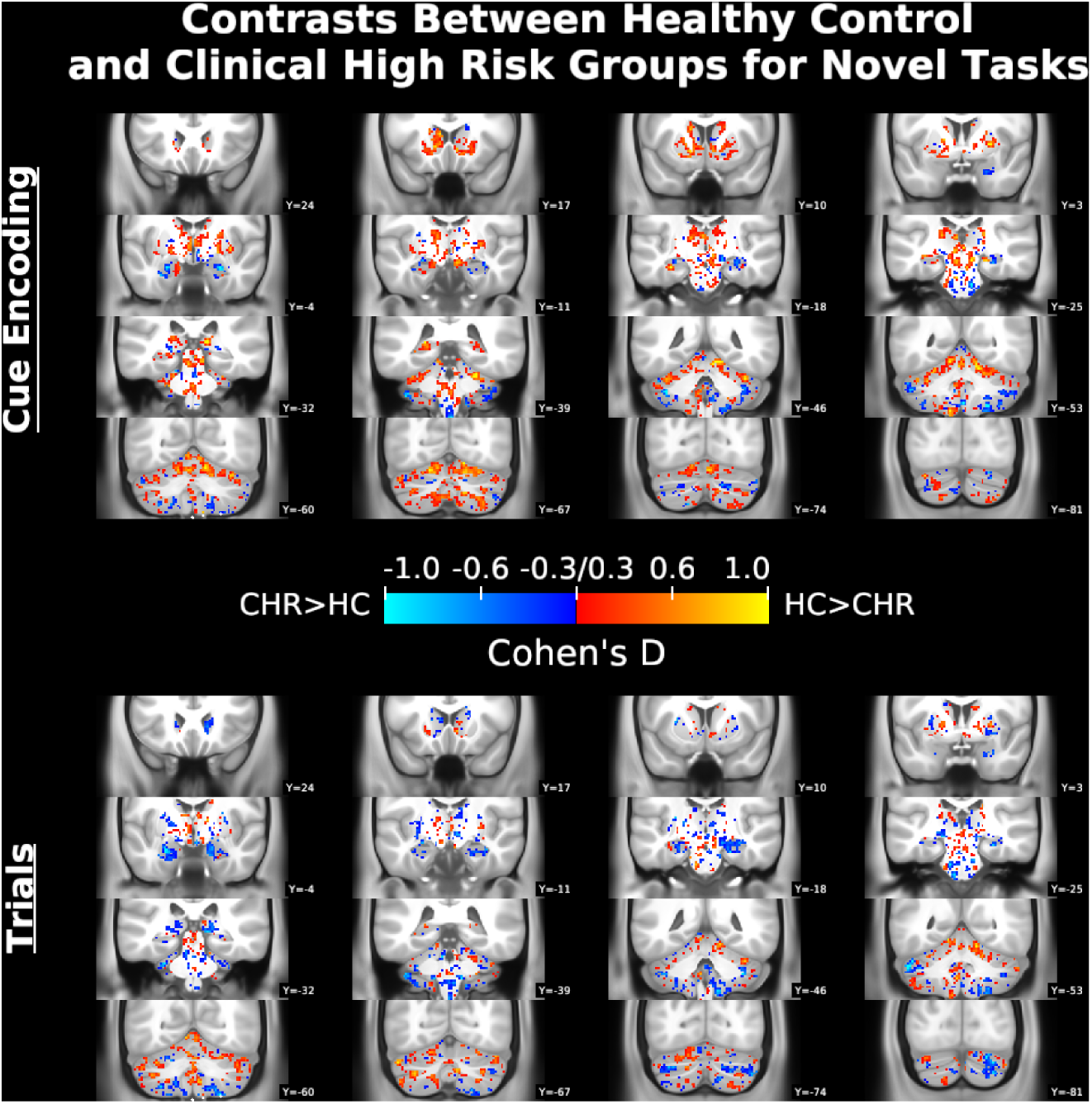
Subcortical effect size maps of contrasts of the Healthy Control (HC) group and the Clinical High Risk (CHR) group for novel tasks at task cue encoding and task performance. The contrast of HC > CHR is shown in hot colors and the contrast of CHR > HC is shown in cold colors. Values shown are Cohen’s D with a minimum threshold of 0.3. Outlines depict the boundaries of the Glasser et al. Multi-modal Parcellation [91], and labels come from that parcellation.

**Supplemental Figure 9.**
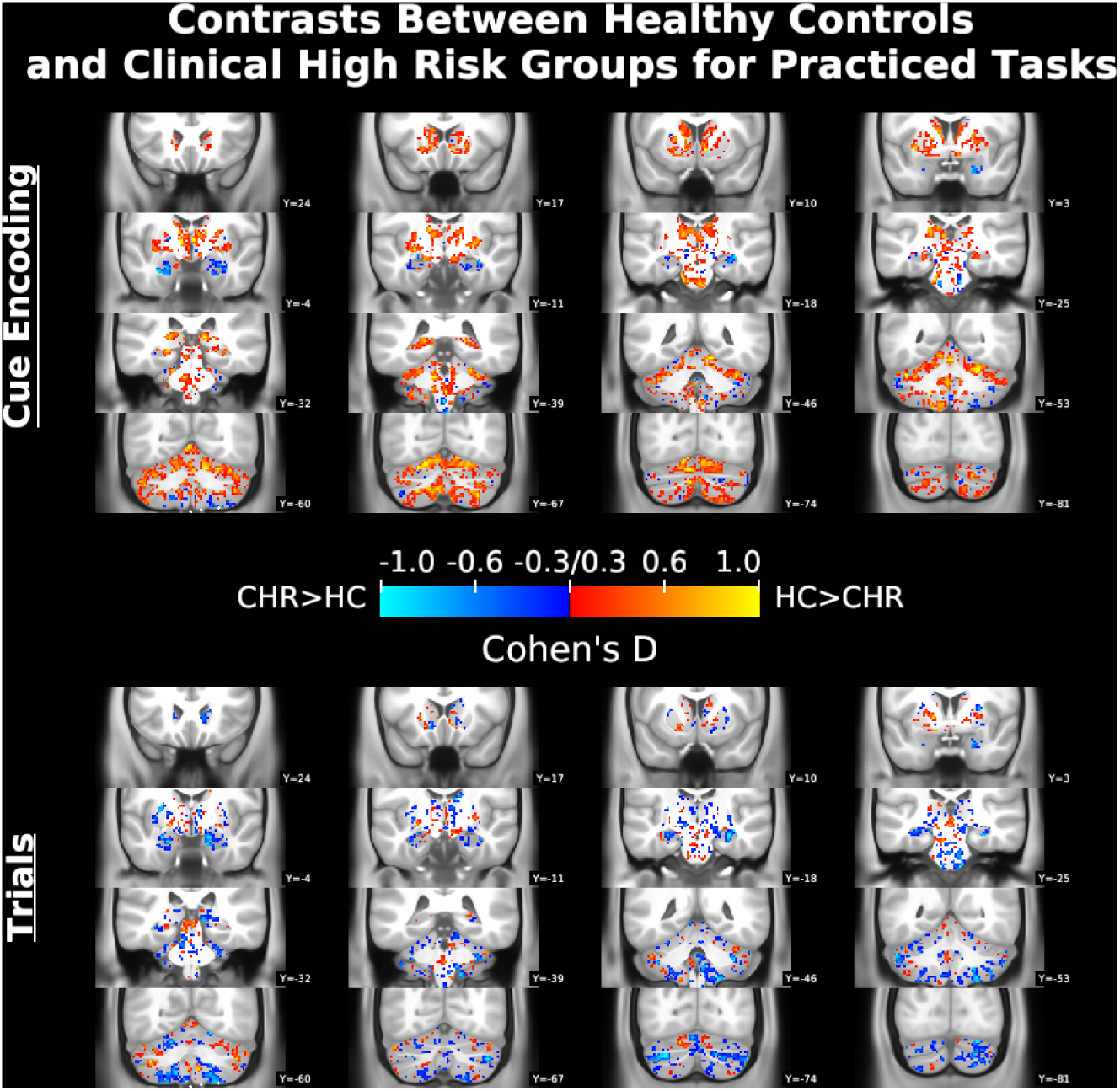
Subcortical effect size maps of contrasts of the Healthy Control (HC) group and the Clinical High Risk (CHR) group for practiced tasks at task cue encoding and task performance. The contrast of HC > CHR is shown in hot colors and the contrast of CHR > HC is shown in cold colors. Values shown are Cohen’s D with a minimum threshold of 0.3. Outlines depict the boundaries of the Glasser et al. Multi-modal Parcellation [91], and labels come from that parcellation.

**Supplemental Figure 10.**
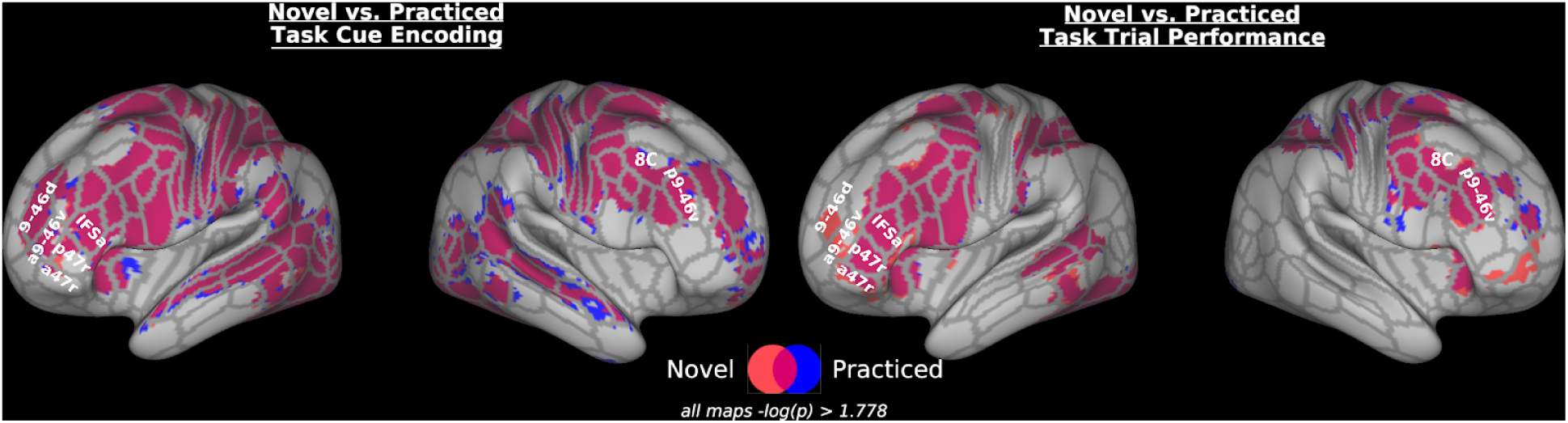
Activation during novel and practiced tasks at task cue encoding and task performance for all participants. Results were corrected with permutation testing and threshold-free cluster enhancement to a threshold of -log(p) > 1.778 (equivalent to p<.05 with Bonferroni correction for 3 tests: Left Cortical Surface, RIght Cortical Surface, and subcortical volume). Novel trial activation is shown in red, practiced task activation in blue, and overlapping activation in purple. Outlines depict the boundaries of the Glasser et al. (2016) Multi-modal Parcellation, and labels come from that parcellation.

**Supplemental Figure 11.**
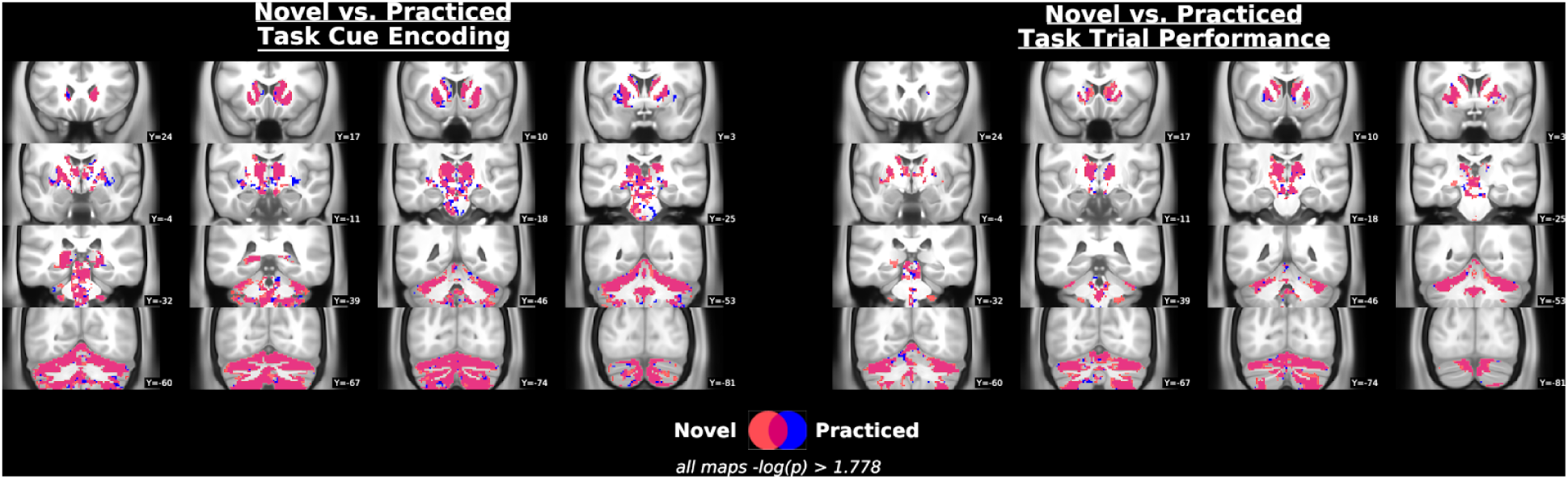
Subcortical activation during novel and practiced tasks at task cue encoding and task performance for all participants. Results were corrected with permutation testing and threshold-free cluster enhancement to a threshold of -log(p) > 1.778 (equivalent to p<.05 with Bonferroni correction for 3 tests: Left Cortical Surface, Right Cortical Surface, and subcortical volume). Novel trial activation is shown in red, practiced task activation in blue, and overlapping activation in purple.

## Footnotes

1 Calculated with G*Power 3.1, as a difference between independent means, 1-tailed.

## References

Abraham, A., Dohmatob, E., Thirion, B., Samaras, D., Varoquaux, G., 2014a. Region segmentation for sparse decompositions: better brain parcellations from rest fMRI. arXiv [q-bio.NC].

Abraham, A., Pedregosa, F., Eickenberg, M., Gervais, P., Mueller, A., Kossaifi, J., Gramfort, A., Thirion, B., Varoquaux, G., 2014b. Machine learning for neuroimaging with scikit-learn. Front. Neuroinform. 8, 14. https://doi.org/10.3389/fninf.2014.00014

Allen, P., Luigjes, J., Howes, O.D., Egerton, A., Hirao, K., Valli, I., Kambeitz, J., Fusar-Poli, P., Broome, M., McGuire, P., 2012. Transition to psychosis associated with prefrontal and subcortical dysfunction in ultra high-risk individuals. Schizophr. Bull. 38, 1268–1276. https://doi.org/10.1093/schbul/sbr194

Amunts, K., Lenzen, M., Friederici, A.D., Schleicher, A., Morosan, P., Palomero-Gallagher, N., Zilles, K., 2010. Broca’s region: novel organizational principles and multiple receptor mapping. PLoS Biol. 8. https://doi.org/10.1371/journal.pbio.1000489

Andersson, J.L.R., Skare, S., Ashburner, J., 2003. How to correct susceptibility distortions in spin-echo echo-planar images: Application to diffusion tensor imaging. Neuroimage 20, 870–888. https://doi.org/10.1016/S1053-8119(03)00336-7

Assem, M., Glasser, M.F., Van Essen, D.C., Duncan, J., 2019. A domain-general cognitive core defined in multimodally parcellated human cortex. BioRxiv.

Avants, B.B., Epstein, C.L., Grossman, M., Gee, J.C., 2008. Symmetric diffeomorphic image registration with cross-correlation: evaluating automated labeling of elderly and neurodegenerative brain. Med. Image Anal. 12, 26–41. https://doi.org/10.1016/j.media.2007.06.004

Badre, D., 2008. Cognitive control, hierarchy, and the rostro–caudal organization of the frontal lobes. Trends Cogn. Sci. 12, 193–200. https://doi.org/10.1016/j.tics.2008.02.004

Badre, D., Nee, D.E., 2017. Frontal Cortex and the Hierarchical Control of Behavior. Trends Cogn. Sci. 22, 170–188. https://doi.org/10.1016/j.tics.2017.11.005

Badre, D., Poldrack, R.A., Paré-Blagoev, E.J., Insler, R.Z., Wagner, A.D., 2005. Dissociable controlled retrieval and generalized selection mechanisms in ventrolateral prefrontal cortex. Neuron 47, 907–918. https://doi.org/10.1016/j.neuron.2005.07.023

Badre, D., Wagner, A.D., 2004. Selection, integration, and conflict monitoring; assessing the nature and generality of prefrontal cognitive control mechanisms. Neuron 41, 473–487.

Barch, D.M., Ceaser, A., 2012. Cognition in schizophrenia: core psychological and neural mechanisms. Trends Cogn. Sci. 16, 27–34. https://doi.org/10.1016/j.tics.2011.11.015

Bode, S., Haynes, J.-D., 2009. Decoding sequential stages of task preparation in the human brain. Neuroimage 45, 606–613. https://doi.org/10.1016/j.neuroimage.2008.11.031

Bora, E., Murray, R.M., 2014. Meta-analysis of Cognitive Deficits in Ultra-high Risk to Psychosis and First-Episode Psychosis: Do the Cognitive Deficits Progress Over, or After, the Onset of Psychosis? Schizophrenia Bulletin. https://doi.org/10.1093/schbul/sbt085

Braver, T.S., Barch, D.M., 2006. Extracting core components of cognitive control. Trends Cogn. Sci. 10, 529–532. https://doi.org/10.1016/j.tics.2006.10.006

Camchong, J., Lim, K.O., Sponheim, S.R., Macdonald, A.W., 2009. Frontal white matter integrity as an endophenotype for schizophrenia: diffusion tensor imaging in monozygotic twins and patients’ nonpsychotic relatives. Front. Hum. Neurosci. 3, 35. https://doi.org/10.3389/neuro.09.035.2009

Cannon, T.D., Cadenhead, K., Cornblatt, B., Woods, S.W., Addington, J., Walker, E., Seidman, L.J., Perkins, D., Tsuang, M., McGlashan, T., Heinnsen, R., 2008. Prediction of Psychosis in youth at High Clinical Risk. Arch. Gen. Psychiatry 65, 28–37.

Cannon, T.D., Yu, C., Addington, J., Bearden, C.E., Cadenhead, K.S., Cornblatt, B.A., Heinssen, R., Jeffries, C.D., Mathalon, D.H., McGlashan, T.H., Perkins, D.O., Seidman, L.J., Tsuang, M.T., Walker, E.F., Woods, S.W., Kattan, M.W., 2016. An individualized risk calculator for research in prodromal psychosis. Am. J. Psychiatry 173, 980–988. https://doi.org/10.1176/appi.ajp.2016.15070890

Carrión, R.E., Walder, D.J., Auther, A.M., McLaughlin, D., Zyla, H.O., Adelsheim, S., Calkins, R., Carter, C.S., McFarland, B., Melton, R., Niendam, T., Ragland, J.D., Sale, T.G., Taylor, S.F., McFarlane, W.R., Cornblatt, B.A., 2018. From the psychosis prodrome to the first-episode of psychosis: No evidence of a cognitive decline. J. Psychiatr. Res. 96, 231–238. https://doi.org/10.1016/j.jpsychires.2017.10.014

Cole, M.W., Bagic, A., Kass, R., Schneider, W., 2010. Prefrontal dynamics underlying rapid instructed task learning reverse with practice. Journal of Neuroscience 30, 14245–14254. https://doi.org/10.1523/JNEUROSCI.1662-10.2010

Cole, M.W., Braver, T.S., Meiran, N., 2017. The task novelty paradox: Flexible control of inflexible neural pathways during rapid instructed task learning. Neurosci. Biobehav. Rev. 81, 4–15. https://doi.org/10.1016/j.neubiorev.2017.02.009

Cole, M.W., Ito, T., Braver, T.S., 2016. The Behavioral Relevance of Task Information in Human Prefrontal Cortex. Cereb. Cortex 1–9. https://doi.org/10.1093/cercor/bhv072

Cole, M.W., Laurent, P., Stocco, A., 2013. Rapid instructed task learning: a new window into the human brain’s unique capacity for flexible cognitive control. Cogn. Affect. Behav. Neurosci. 13, 1–22. https://doi.org/10.3758/s13415-012-0125-7

Cox, R.W., 1996. AFNI: Software for analysis and visualization of functional magnetic resonance neuroimages. Comput. Biomed. Res. 29, 162–173.

Crittenden, B.M., Duncan, J., 2012. Task difficulty manipulation reveals multiple demand activity but no frontal lobe hierarchy. Cereb. Cortex 24, 532–540.

Crone, E.A., Wendelken, C., Donohue, S.E., Bunge, S.A., 2005. Neural evidence for dissociable components of task-switching. Cereb. Cortex 16, 475–486.

Dale, A.M., Fischl, B., Sereno, M.I., 1999. Cortical surface-based analysis. I. Segmentation and surface reconstruction. Neuroimage 9, 179–194. https://doi.org/10.1006/nimg.1998.0395

Damme, K.S.F., Pelletier-Baldelli, A., Cowan, H.R., Orr, J.M., Mittal, V.A., 2019. Distinct and opposite profiles of connectivity during self-reference task and rest in youth at clinical high risk for psychosis. Hum. Brain Mapp. hbm.24595. https://doi.org/10.1002/hbm.24595

Dickie, E.W., Anticevic, A., Smith, D.E., Coalson, T.S., Manogaran, M., Calarco, N., Viviano, J.D., Glasser, M.F., Van Essen, D.C., Voineskos, A.N., 2019. Ciftify: A framework for surface-based analysis of legacy MR acquisitions. Neuroimage 197, 818–826. https://doi.org/10.1016/j.neuroimage.2019.04.078

Donohue, S.E., Wendelken, C., Crone, E.A., Bunge, S.A., 2005. Retrieving rules for behavior from long-term memory. NeuroImage. https://doi.org/10.1016/j.neuroimage.2005.03.019

Dumontheil, I., Thompson, R., Duncan, J., 2011. Assembly and use of new task rules in fronto-parietal cortex. J. Cogn. Neurosci. 23, 168–182. https://doi.org/10.1162/jocn.2010.21439

Duncan, J., 2010. The multiple-demand (MD) system of the primate brain: mental programs for intelligent behaviour. Trends Cogn. Sci. 14, 172–179. https://doi.org/10.1016/j.tics.2010.01.004

Duncan, J., 1986. Disorganisation of behavior after frontal lobe damage. Cogn. Neuropsychol. 3, 271–290.

Esteban, O., Markiewicz, C.J., Blair, R.W., Moodie, C.A., Isik, A.I., Erramuzpe, A., Kent, J.D., Goncalves, M., DuPre, E., Snyder, M., Oya, H., Ghosh, S.S., Wright, J., Durnez, J., Poldrack, R.A., Gorgolewski, K.J., 2019. fMRIPrep: a robust preprocessing pipeline for functional MRI. Nat. Methods 16, 111–116. https://doi.org/10.1038/s41592-018-0235-4

Fassbender, C., Scangos, K., Lesh, T.A., Carter, C.S., 2014. RT distributional analysis of cognitive-control-related brain activity in first-episode schizophrenia. Cogn. Affect. Behav. Neurosci. 14, 175–188. https://doi.org/10.3758/s13415-014-0252-4

First, M., Spitzer, R., Gibbon, M., Williams, J., 1995. Structured Clinical Interview for the DSM-IV Axis I Disorders (SCID-I), Patient Edition. American Psychiatric Press, Washington DC.

Fonov, V.S., Evans, A.C., McKinstry, R.C., Almli, C.R., Collins, D.L., 2009. Unbiased nonlinear average age-appropriate brain templates from birth to adulthood. Neuroimage 47, S102. https://doi.org/10.1016/S1053-8119(09)70884-5

Fornito, A., Harrison, B.J., Goodby, E., Dean, A., Ooi, C., Nathan, P.J., Lennox, B.R., Jones, P.B., Suckling, J., Bullmore, E.T., 2013. Functional dysconnectivity of corticostriatal circuitry as a risk phenotype for psychosis. JAMA Psychiatry 70, 1143–1151. https://doi.org/10.1001/jamapsychiatry.2013.1976

Fusar-Poli, P., Cappucciati, M., Borgwardt, S., Woods, S.W., Addington, J., Nelson, B., Nieman, D.H., Stahl, D.R., Rutigliano, G., Riecher-Rössler, A., Simon, A.E., Mizuno, M., Lee, T.Y., Kwon, J.S., Lam, M.M.L., Perez, J., Keri, S., Amminger, P., Metzler, S., Kawohl, W., Rössler, W., Lee, J., Labad, J., Ziermans, T., An, S.K., Liu, C.-C., Woodberry, K.A., Braham, A., Corcoran, C., McGorry, P., Yung, A.R., McGuire, P.K., 2016. Heterogeneity of Psychosis Risk Within Individuals at Clinical High Risk: A Meta-analytical Stratification. JAMA Psychiatry 73, 113–120. https://doi.org/10.1001/jamapsychiatry.2015.2324

Fusar-Poli, P., Deste, G., Smieskova, R., Barlati, S., Yung, A.R., Howes, O., Stieglitz, R.-D., Vita, A., McGuire, P., Borgwardt, S., 2012. Cognitive functioning in prodromal psychosis: a meta-analysis. Arch. Gen. Psychiatry 69, 562–571. https://doi.org/10.1001/archgenpsychiatry.2011.1592

Fusar-Poli, P., Howes, O.D., Allen, P., Broome, M., Valli, I., Asselin, M.-C., Montgomery, A.J., Grasby, P.M., McGuire, P., 2011. Abnormal prefrontal activation directly related to pre-synaptic striatal dopamine dysfunction in people at clinical high risk for psychosis. Mol. Psychiatry 16, 67–75. https://doi.org/10.1038/mp.2009.108

Glasser, M.F., Coalson, T.S., Robinson, E.C., Hacker, C.D., Harwell, J., Yacoub, E., Ugurbil, K., Andersson, J., Beckmann, C.F., Jenkinson, M., Smith, S.M., Van Essen, D.C., 2016. A multi-modal parcellation of human cerebral cortex. Nature 536, 171–178. https://doi.org/10.1038/nature18933

Glasser, M.F., Sotiropoulos, S.N., Wilson, J.A., Coalson, T.S., Fischl, B., Andersson, J.L.R., Xu, J., Jbabdi, S., Webster, M., Polimeni, J.R., Van Essen, D.C., Jenkinson, M., 2013. The minimal preprocessing pipelines for the human connectome project. Neuroimage 80, 105–124. https://doi.org/10.1016/j.neuroimage.2013.04.127

Gorgolewski, K.J., Auer, T., Calhoun, V.D., Craddock, R.C., Das, S., Duff, E.P., Flandin, G., Ghosh, S.S., Glatard, T., Halchenko, Y.O., Handwerker, D.A., Hanke, M., Keator, D., Li, X., Michael, Z., Maumet, C., Nichols, B.N., Nichols, T.E., Pellman, J., Poline, J.-B., Rokem, A., Schaefer, G., Sochat, V., Triplett, W., Turner, J.A., Varoquaux, G., Poldrack, R.A., 2016. The brain imaging data structure, a format for organizing and describing outputs of neuroimaging experiments. Sci Data 3, 160044. https://doi.org/10.1038/sdata.2016.44

Gorgolewski, K.J., Burns, C.D., Madison, C., Clark, D., Halchenko, Y.O., Waskom, M.L., Ghosh, S.S., 2011. Nipype: A Flexible, Lightweight and Extensible Neuroimaging Data Processing Framework in Python. Front. Neuroinform. 5, 13. https://doi.org/10.3389/fninf.2011.00013

Gorgolewski, K.J., Esteban, O., Ellis, D.G., Notter, M.P., Ziegler, E., Johnson, H., Hamalainen, C., Yvernault, B., Burns, C., Manhães-Savio, A., Jarecka, D., Markiewicz, C.J., Salo, T., Clark, D., Waskom, M., Wong, J., Modat, M., Dewey, B.E., Clark, M.G., Dayan, M., Loney, F., Madison, C., Gramfort, A., Keshavan, A., Berleant, S., Pinsard, B., Goncalves, M., Clark, D., Cipollini, B., Varoquaux, G., Wassermann, D., Rokem, A., Halchenko, Y.O., Forbes, J., Moloney, B., Malone, I.B., Hanke, M., Mordom, D., Buchanan, C., Pauli, W.M., Huntenburg, J.M., Horea, C., Schwartz, Y., Tungaraza, R., Iqbal, S., Kleesiek, J., Sikka, S., Frohlich, C., Kent, J., Perez-Guevara, M., Watanabe, A., Welch, D., Cumba, C., Ginsburg, D., Eshaghi, A., Kastman, E., Bougacha, S., Blair, R., Acland, B., Gillman, A., Schaefer, A., Nichols, B.N., Giavasis, S., Erickson, D., Correa, C., Ghayoor, A., Küttner, R., Haselgrove, C., Zhou, D., Craddock, R.C., Haehn, D., Lampe, L., Millman, J., Lai, J., Renfro, M., Liu, S., Stadler, J., Glatard, T., Kahn, A.E., Kong, X.-Z., Triplett, W., Park, A., McDermottroe, C., Hallquist, M., Poldrack, R.A., Perkins, L.N., Noel, M., Gerhard, S., Salvatore, J., Mertz, F., Broderick, W., Inati, S., Hinds, O., Brett, M., Durnez, J., Tambini, A., Rothmei, S., Andberg, S.K., Cooper, G., Marina, A., Mattfeld, A., Urchs, S., Sharp, P., Matsubara, K., Geisler, D., Cheung, B., Floren, A., Nickson, T., Pannetier, N., Weinstein, A., Dubois, M., Arias, J., Tarbert, C., Schlamp, K., Jordan, K., Liem, F., Saase, V., Harms, R., Khanuja, R., Podranski, K., Flandin, G., Papadopoulos Orfanos, D., Schwabacher, I., McNamee, D., Falkiewicz, M., Pellman, J., Linkersdörfer, J., Varada, J., Pérez-García, F., Davison, A., Shachnev, D., Ghosh, S., 2017. Nipype: a flexible, lightweight and extensible neuroimaging data processing framework in Python. 0.13.1. https://doi.org/10.5281/ZENODO.581704

Greve, D.N., Fischl, B., 2009. Accurate and robust brain image alignment using boundary-based registration. Neuroimage 48, 63–72. https://doi.org/10.1016/j.neuroimage.2009.06.060

Guo, J.Y., Niendam, T.A., Auther, A.M., Carrión, R.E., Cornblatt, B.A., Ragland, J.D., Adelsheim, S., Calkins, R., Sale, T.G., Taylor, S.F., McFarlane, W.R., Carter, C.S., 2019. Predicting psychosis risk using a specific measure of cognitive control: a 12-month longitudinal study. Psychol. Med. 1–10. https://doi.org/10.1017/S0033291719002332

Gur, R.E., Cowell, P.E., Latshaw, A., Turetsky, B.I., Grossman, R.I., Arnold, S.E., Bilker, W.B., Gur, R.C., 2000. Reduced dorsal and orbital prefrontal gray matter volumes in schizophrenia. Arch. Gen. Psychiatry 57, 761–768.

Haroun, N., Dunn, L., Haroun, A., Cadenhead, K.S., 2006. Risk and protection in prodromal schizophrenia: ethical implications for clinical practice and future research. Schizophr. Bull. 32, 166–178. https://doi.org/10.1093/schbul/sbj007

Harris, J.M., Whalley, H., Yates, S., Miller, P., Johnstone, E.C., Lawrie, S.M., 2004. Abnormal cortical folding in high-risk individuals: A predictor of the development of schizophrenia? Biol. Psychiatry 56, 182–189. https://doi.org/10.1016/j.biopsych.2004.04.007

Jenkinson, M., Bannister, P., Brady, M., Smith, S.M., 2002. Improved Optimization for the Robust and Accurate Linear Registration and Motion Correction of Brain Images. Neuroimage 17, 825–841. https://doi.org/10.1006/nimg.2002.1132

Kim, C., Cilles, S.E., Johnson, N.F., Gold, B.T., 2012. Domain general and domain preferential brain regions associated with different types of task switching: A Meta-Analysis. Hum. Brain Mapp. 33, 130–142. https://doi.org/10.1002/hbm.21199

Kostopoulos, P., Petrides, M., 2003. The mid-ventrolateral prefrontal cortex: insights into its role in memory retrieval. Eur. J. Neurosci. 17, 1489–1497.

Lesh, T.A., Westphal, A.J., Niendam, T.A., Yoon, J.H., Minzenberg, M.J., Ragland, J.D., Solomon, M., Carter, C.S., 2013. Proactive and reactive cognitive control and dorsolateral prefrontal cortex dysfunction in first episode schizophrenia. Neuroimage Clin 2, 590–599. https://doi.org/10.1016/j.nicl.2013.04.010

MacDonald, A.W., 3rd, Carter, C.S., 2003. Event-related FMRI study of context processing in dorsolateral prefrontal cortex of patients with schizophrenia. J. Abnorm. Psychol. 112, 689–697. https://doi.org/10.1037/0021-843X.112.4.689

MacDonald, A.W., 3rd, Carter, C.S., Kerns, J.G., Ursu, S., Barch, D.M., Holmes, A.J., Stenger, V.A., Cohen, J.D., 2005. Specificity of prefrontal dysfunction and context processing deficits to schizophrenia in never-medicated patients with first-episode psychosis. Am. J. Psychiatry 162, 475–484. https://doi.org/10.1176/appi.ajp.162.3.475

Mann, C.L., Footer, O., Chung, Y.S., Driscoll, L.L., Barch, D.M., 2013. Spared and impaired aspects of motivated cognitive control in schizophrenia. J. Abnorm. Psychol. 122, 745–755. https://doi.org/10.1037/a0033069

McGlashan, T.H., Addington, J., Cannon, T., 2007. Recruitment and treatment practices for help-seeking “prodromal” patients. Schizophrenia.

Miller, E.K., Cohen, J.D., 2001. An integrative theory of prefrontal cortex function. Annu. Rev. Neurosci. 24, 167–202. https://doi.org/10.1146/annurev.neuro.24.1.167

Miller, T.J., Mcglashan, T.H., Woods, S.W., Stein, K., Driesen, N., Corcoran, C.M., Hoffman, R., Davidson, L., 1999. Symptom assessment in schizophrenic prodromal states. Psychiatr. Q. 70, 273–287.

Minzenberg, M.J., Laird, A.R., Thelen, S., Carter, C.S., Glahn, D.C., 2009. Meta-analysis of 41 functional neuroimaging studies of executive function in schizophrenia. Arch. Gen. Psychiatry 66, 811–822. https://doi.org/10.1001/archgenpsychiatry.2009.91

Monsell, S., 1996. Control of mental processes. Unsolved mysteries of the mind: Tutorial essays in cognition 93–148.

Morey, R.A., Inan, S., Mitchell, T.V., Perkins, D.O., Lieberman, J.A., Belger, A., 2005. Imaging frontostriatal function in ultra-high-risk, early, and chronic schizophrenia during executive processing. Arch. Gen. Psychiatry 62, 254–262. https://doi.org/10.1001/archpsyc.62.3.254

Neubert, F.-X., Mars, R.B., Thomas, A.G., Sallet, J., Rushworth, M.F.S., 2014. Comparison of human ventral frontal cortex areas for cognitive control and language with areas in monkey frontal cortex. Neuron 81, 700–713. https://doi.org/10.1016/j.neuron.2013.11.012

Niendam, T.A., Lesh, T.A., Yoon, J., Westphal, A.J., Hutchinson, N., Ragland, J.D., Solomon, M., Minzenberg, M., Carter, C.S., 2014. Impaired context processing as a potential marker of psychosis risk state | Kopernio. Psychiatry Res. 221, 13–20. https://doi.org/10.1016/j.pscychresns.2013.09.001

Oh, J.S., Kubicki, M., Rosenberger, G., Bouix, S., Levitt, J.J., McCarley, R.W., Westin, C.-F., Shenton, M.E., 2009. Thalamo-frontal white matter alterations in chronic schizophrenia: a quantitative diffusion tractography study. Hum. Brain Mapp. 30, 3812–3825. https://doi.org/10.1002/hbm.20809

Pelletier-Baldelli, A., Orr, J.M., Bernard, J.A., Mittal, V.A., 2018. Social reward processing: A biomarker for predicting psychosis risk? Schizophr. Res. https://doi.org/10.1016/j.schres.2018.07.042

Pettersson-Yeo, W., Allen, P., Benetti, S., McGuire, P., Mechelli, A., 2011. Dysconnectivity in schizophrenia: where are we now? Neurosci. Biobehav. Rev. 35, 1110–1124. https://doi.org/10.1016/j.neubiorev.2010.11.004

Poppe, A.B., Barch, D.M., Carter, C.S., Gold, J.M., Ragland, J.D., Silverstein, S.M., MacDonald, A.W., 2016. Reduced Frontoparietal Activity in Schizophrenia Is Linked to a Specific Deficit in Goal Maintenance: A Multisite Functional Imaging Study. Schizophr. Bull. 1–9. https://doi.org/10.1093/schbul/sbw036

Pruim, R.H.R., Mennes, M., van Rooij, D., Llera, A., Buitelaar, J.K., Beckmann, C.F., 2015. ICA-AROMA: A robust ICA-based strategy for removing motion artifacts from fMRI data. Neuroimage 112, 267–277. https://doi.org/10.1016/j.neuroimage.2015.02.064

Ragland, J.D., Blumenfeld, R.S., Ramsay, I.S., Yonelinas, A., Yoon, J., Solomon, M., Carter, C.S., Ranganath, C., 2012. Neural correlates of relational and item-specific encoding during working and long-term memory in schizophrenia. Neuroimage 59, 1719–1726. https://doi.org/10.1016/j.neuroimage.2011.08.055

Repovs, G., Csernansky, J.G., Barch, D.M., 2011. Brain network connectivity in individuals with schizophrenia and their siblings. Biol. Psychiatry 69, 967–973. https://doi.org/10.1016/j.biopsych.2010.11.009

Robinson, E.C., Garcia, K., Glasser, M.F., Chen, Z., Coalson, T.S., Makropoulos, A., Bozek, J., Wright, R., Schuh, A., Webster, M., Hutter, J., Price, A., Cordero Grande, L., Hughes, E., Tusor, N., Bayly, P.V., Van Essen, D.C., Smith, S.M., Edwards, A.D., Hajnal, J., Jenkinson, M., Glocker, B., Rueckert, D., 2018. Multimodal surface matching with higher-order smoothness constraints. Neuroimage 167, 453–465. https://doi.org/10.1016/j.neuroimage.2017.10.037

Robinson, E.C., Jbabdi, S., Glasser, M.F., Andersson, J., Burgess, G.C., Harms, M.P., Smith, S.M., Van Essen, D.C., Jenkinson, M., 2014. MSM: a new flexible framework for Multimodal Surface Matching. Neuroimage 100, 414–426. https://doi.org/10.1016/j.neuroimage.2014.05.069

Rotarska-Jagiela, A., van de Ven, V., Oertel-Knöchel, V., Uhlhaas, P.J., Vogeley, K., Linden, D.E.J., 2010. Resting-state functional network correlates of psychotic symptoms in schizophrenia. Schizophr. Res. 117, 21–30. https://doi.org/10.1016/j.schres.2010.01.001

Ruge, H., Jamadar, S., Zimmermann, U., Karayanidis, F., 2013. The many faces of preparatory control in task switching: reviewing a decade of fMRI research. Hum. Brain Mapp. 34, 12–35. https://doi.org/10.1002/hbm.21420

Sallet, P.C., Elkis, H., Alves, T.M., Oliveira, J.R., Sassi, E., Campi de Castro, C., Busatto, G.F., Gattaz, W.F., 2003. Reduced cortical folding in schizophrenia: an MRI morphometric study. Am. J. Psychiatry 160, 1606–1613. https://doi.org/10.1176/appi.ajp.160.9.1606

Seidman, L.J., Thermenos, H.W., Poldrack, R.A., Peace, N.K., Koch, J.K., Faraone, S.V., Tsuang, M.T., 2006. Altered brain activation in dorsolateral prefrontal cortex in adolescents and young adults at genetic risk for schizophrenia: an fMRI study of working memory. Schizophr. Res. 85, 58–72. https://doi.org/10.1016/j.schres.2006.03.019

Sheffield, J.M., Gold, J.M., Strauss, M.E., Carter, C.S., MacDonald, A.W., Ragland, J.D., Silverstein, S.M., Barch, D.M., 2014. Common and specific cognitive deficits in schizophrenia: relationships to function. Cogn. Affect. Behav. Neurosci. 14, 161–174. https://doi.org/10.3758/s13415-013-0211-5

Smith, S.M., Jenkinson, M., Woolrich, M.W., Beckmann, C.F., Behrens, T.E.J., Johansen-Berg, H., Bannister, P.R., De Luca, M., Drobnjak, I., Flitney, D.E., Niazy, R.K., Saunders, J., Vickers, J., Zhang, Y., De Stefano, N., Brady, J.M., Matthews, P.M., 2004. Advances in functional and structural MR image analysis and implementation as FSL. Neuroimage 23 Suppl 1, S208–S219. https://doi.org/10.1016/j.neuroimage.2004.07.051

Stanfield, A.C., Moorhead, T.W.J., Harris, J.M., Owens, D.G.C., Lawrie, S.M., Johnstone, E.C., 2008. Increased Right Prefrontal Cortical Folding in Adolescents at Risk of Schizophrenia for Cognitive Reasons. Biol. Psychiatry 63, 80–85. https://doi.org/10.1016/j.biopsych.2007.04.012

Stuss, D.T., Alexander, M.P., 2000. Executive functions and the frontal lobes: a conceptual view. Psychol. Res. 63, 289–298. https://doi.org/10.1007/s004269900007. The jamovi project, 2019. jamovi

Thompson, E., Millman, Z.B., Okuzawa, N., Mittal, V., DeVylder, J., Skadberg, T., Buchanan, R.W., Reeves, G.M., Schiffman, J., 2015. Evidence-based early interventions for individuals at clinical high risk for psychosis: a review of treatment components. J. Nerv. Ment. Dis. 203, 342–351. https://doi.org/10.1097/NMD.0000000000000287

Tustison, N.J., Avants, B.B., Cook, P.A., Yuanjie Zheng, Egan, A., Yushkevich, P.A., Gee, J.C., 2010. N4ITK: Improved N3 Bias Correction. IEEE Trans. Med. Imaging 29, 1310–1320. https://doi.org/10.1109/TMI.2010.2046908

van den Heuvel, M.P., Mandl, R.C.W., Stam, C.J., Kahn, R.S., Pol, H.E.H., 2010. Aberrant Frontal and Temporal Complex Network Structure in Schizophrenia: A Graph Theoretical Analysis. Journal of Neuroscience 30, 15915–15926. https://doi.org/10.1523/JNEUROSCI.2874-10.2010

Winkler, A.M., Ridgway, G.R., Webster, M.A., Smith, S.M., Nichols, T.E., 2014. Permutation inference for the general linear model. Neuroimage 92, 381–397. https://doi.org/10.1016/j.neuroimage.2014.01.060

Woods, S.W., Walsh, B.C., Addington, J., Cadenhead, K.S., Cannon, T.D., Cornblatt, B.A., Heinssen, R., Perkins, D.O., Seidman, L.J., Tarbox, S.I., Tsuang, M.T., Walker, E.F., McGlashan, T.H., 2014. Current status specifiers for patients at clinical high risk for psychosis. Schizophr. Res. 158, 69–75. https://doi.org/10.1016/j.schres.2014.06.022

Woolgar, A., Hampshire, A., Thompson, R., Duncan, J., 2011. Adaptive coding of task-relevant information in human frontoparietal cortex. Journal of Neuroscience 31, 14592–14599. https://doi.org/10.1523/JNEUROSCI.2616-11.2011

Woolrich, M.W., Ripley, B.D., Brady, M., Smith, S.M., 2001. Temporal Autocorrelation in Univariate Linear Modeling of FMRI Data. Neuroimage 14, 1370–1386. https://doi.org/10.1006/nimg.2001.0931

Yung, A.R., McGorry, P.D., 1996. The prodromal phase of first-episode psychosis: past and current conceptualizations. Schizophr. Bull. 22, 353–370. https://doi.org/10.1093/schbul/22.2.353

Yung, A.R., Yuen, H.P., Berger, G., Francey, S., Hung, T.-C., Nelson, B., Phillips, L., McGorry, P., 2007. Declining transition rate in ultra high risk (prodromal) services: dilution or reduction of risk? Schizophr. Bull. 33, 673–681. https://doi.org/10.1093/schbul/sbm015

Zhang, Y., Brady, M., Smith, S., 2001. Segmentation of brain MR images through a hidden Markov random field model and the expectation-maximization algorithm. IEEE Trans. Med. Imaging 20, 45–57. https://doi.org/10.1109/42.906424

Zhou, Y., Liang, M., Jiang, T., Tian, L., Liu, Y., Liu, Z., Liu, H., Kuang, F., 2007. Functional dysconnectivity of the dorsolateral prefrontal cortex in first-episode schizophrenia using resting-state fMRI. Neurosci. Lett. 417, 297–302. https://doi.org/10.1016/j.neulet.2007.02.081

